# Mapping of the gene network that regulates glycan clock of ageing

**DOI:** 10.1101/2023.04.25.23289027

**Authors:** Azra Frkatović-Hodžić, Karlo Miškec, Anika Mijakovac, Arina Nostaeva, Sodbo Z. Sharapov, Arianna Landini, Toomas Haller, Erik van den Akker, Sapna Sharma, Rafael R. C. Cuadrat, Massimo Mangino, Yong Li, Toma Keser, Najda Rudman, Tamara Štambuk, Maja Pučić-Baković, Irena Trbojević-Akmačić, Ivan Gudelj, Jerko Štambuk, Tea Pribić, Barbara Radovani, Petra Tominac, Krista Fischer, Marian Beekman, Manfred Wuhrer, Christian Gieger, Matthias B. Schulze, Clemens Wittenbecher, Ozren Polasek, Caroline Hayward, James F. Wilson, Tim D. Spector, Anna Köttgen, Frano Vučković, Yurii S. Aulchenko, Aleksandar Vojta, Jasminka Krištić, Lucija Klarić, Vlatka Zoldoš, Gordan Lauc

## Abstract

Glycans are an essential structural component of Immunoglobulin G (IgG) that modulate its structure and function. However, regulatory mechanisms behind this complex posttranslational modification are not well known. Previous genome-wide association studies (GWAS) identified 29 genomic regions involved in regulation of IgG glycosylation, but only a few were functionally validated. One of the key functional features of IgG glycosylation is the addition of galactose (galactosylation). We performed GWAS of IgG galactosylation (N=13,705) and identified 16 significantly associated loci, indicating that IgG galactosylation is regulated by a complex network of genes that extends beyond the galactosyltransferase enzyme that adds galactose to IgG glycans. Gene prioritization identified 37 candidate genes. Using a recently developed CRISPR/dCas9 system we manipulated gene expression of candidate genes in the *in vitro* IgG expression system. Up- and downregulation of three genes, *EEF1A1, MANBA* and *TNFRSF13B*, changed the IgG glycome composition, which confirmed that these three genes are involved in IgG galactosylation in this *in vitro* expression system.

## Introduction

Glycosylation is a posttranslational modification characterized by the attachment of oligosaccharide chains (glycans) to proteins or lipids^1^. Glycans linked to immunoglobulin G (IgG) are essential regulators of effector functions of both native and therapeutic monoclonal antibodies. At the population level, galactosylation (the presence of zero (G0), one (G1), or two (G2) galactose moieties) is the most variable trait in the human IgG glycome^2,3^, with agalactosylated glycans comprising 6%-50% of the total IgG glycome in healthy subjects^4^. Galactosylation is also the most variable glycan trait between different strains of mice^5^. The presence of galactose on IgG glycans has an important role in the downstream immune responses mediated by IgG antibodies. Most of the studies show that galactosylated IgG has increased complement and FcγR activity^6–10^. van Osch *et al*.^8^ even proposed a mechanism where IgG galactosylation promotes the hexamerization of IgG antibodies thus enhancing the activation of the classical complement pathway.

Large-scale studies have shown that IgG galactosylation levels decrease with ageing and in inflammatory conditions^11^ and enabled the development of the glycan clock of ageing^12^. The IgG glycome composition has been found to associate with age with galactosylation reaching a peak in early adulthood and then declining with age^13,14^. It was proposed that agalactosylated IgG which increases with age acts as an effector of pro-inflammatory pathological changes and therefore can be exploited not only as a biomarker, but also functional effector of ageing^15,16^.

Elevated levels of agalactosylated IgG glycans were first observed in rheumatoid arthritis ^17^, and followed by similar discoveries in various autoimmune conditions, such as systemic lupus erythematosus (SLE), inflammatory bowel disease, active spondyloarthropathy, autoimmune vasculitis and adult periodontal disease^18–22^. Changes in levels of galactosylated IgG also occur in infectious diseases such as COVID-19^23^. In addition, decreased IgG galactosylation levels were observed in cancer patients (for a review see ^11^ and references herein), potentially reflecting the defensive inflammatory response to cancer^24^ and acute-phase response processes involved in cancer progression^25^. However, the question remains whether the agalactosylated IgG is just a biomarker, or whether it can functionally contribute to disease activity.

Several studies have demonstrated that the regulation of IgG glycosylation is largely influenced by genetics. Up to 75% of the variance in some IgG glycan traits can be explained by the genetic component^26^. The genetic determinants of populational variation in IgG glycosylation were explored by genome-wide association studies (GWAS) which identified at least 29 candidate genomic loci^27–31^. However, the involvement of the specific genes and their potential interactions to generate differential IgG galactosylation are still poorly understood. Here, we conducted a GWAS of IgG galactosylation phenotypes in a study that almost doubles the sample size (N=13,705) compared to previous GWAS of IgG N-glycome^30^ and focus on the genes with *in-silico* evidence for involvement in the IgG galactosylation process. To assess their functional role in IgG galactosylation we applied our recently developed HEK293FS IgG transient expression system based on CRISPR-dCas9 molecular tools to functionally test whether the change in the expression of associated genes affects the IgG galactosylation levels^32^.

## Results

To enable meta-analysis of GWAS of galactosylation phenotypes measured by different analytical platforms and thereby increase the total number of samples, we first developed a protocol for data harmonisation for glycan data generated using different analytical platforms (UPLC and LC-MS). The protocol was established based on the estimated correlation of the glycan traits derived in one cohort that had glycans measured using both UPLC and LC-MS platforms. The aim was to combine IgG subclass-specific glycan information obtained from LC-MS (i.e. galactosylation measured separately on IgG1-4 subclasses) in an appropriate manner to obtain information corresponding to the total IgG galactosylation measured by UPLC (analysis of glycans from both Fab and Fc fragments from all IgG subclasses). As described in Supplementary Methods, we compared different normalization and subclass information summary approaches. In a subset of individuals that were analysed using both methods, the Pearson correlation coefficients ranged from 0.95 and 0.97 for G0, 0.73 to 0.80 for G1, and 0.90 and 0.91 for G2 trait (Appendix Table 1). Different pre-processing approaches performed similarly, so we selected median quotient normalization due to previous recommendation^33^, followed by the calculation of the derived traits without IgG subclass weighting in LC-MS data.

**Table 1:** List of genome-wide significant loci in IgG galactosylation GWAS. Locus- chromosome:start- end position in GRCh37 (hg19) build; Top SNP- rsID identifier for the SNP with the strongest association with the glycan trait; Trait-trait with the lowest p-value for the top SNP in the genomic locus; EA- effect allele for which the effect is reported; OA- non-effect allele; EAF- effect allele frequency; Beta- effect estimate for the effect allele of the top SNP; SE- standard error of the effect estimate; P– p-value for the top SNP-trait association; Beta repl- effect estimate for the effect allele of the top SNP in replication analysis; SE repl-standard error of the effect estimate in the replication analysis; P repl- p-value for the top SNP in the replication analysis; Closest gene- gene found closest to the top SNP in the locus; Prioritized genes-symbol for genes prioritized in the locus;

| Locus | Top SNP | Trait | EA | OA | EAF | Beta | SE | P | Beta repl | SE repl | P repl | Closest gene | Prioritized.genes |
| --- | --- | --- | --- | --- | --- | --- | --- | --- | --- | --- | --- | --- | --- |
| 1:24699711-25493756 | rs188468174 | G0 | T | C | 0.015 | 0.693 | 0.048 | 3.20E-47 | 0.865 | 0.076 | 2.96E-30 | *RUNX3* | *RUNX3* |
| 2:26109539-26149988 | rs10177977 | G0 | T | C | 0.334 | -0.062 | 0.01 | 3.80E-10 | -0.049 | 0.015 | 1.03E-03 | *KIF3C* | *KIF3C* |
| 4:103390496-103567348 | rs3774964 | G2 | A | G | 0.37 | -0.065 | 0.01 | 1.56E-11 | -0.038 | 0.015 | 9.20E-03 | *NFKB1* | *NFKB1; MANBA* |
| 6:31107733-31164511 | rs1265109 | G0 | T | G | 0.295 | -0.063 | 0.011 | 1.57E-09 | 0.013 | 0.019 | 4.97E-01 | *HLA region* | *HLA region* |
| 6:74168723-74285118 | rs3822960 | G2 | T | C | 0.306 | 0.061 | 0.01 | 5.84E-10 | 0.028 | 0.015 | 6.24E-02 | *EEF1A1* | *EEF1A1; MTO1* |
| 6:143088071-143206826 | rs7758383 | G2 | A | G | 0.486 | 0.092 | 0.009 | 6.09E-23 | 0.145 | 0.014 | 9.44E-25 | *HIVEP2* | *HIVEP2* |
| 7:150856165-150906453 | rs113745074 | G0 | T | C | 0.094 | 0.12 | 0.015 | 6.19E-16 | 0.134 | 0.022 | 8.47E-10 | *SMARCD3* | *ABCF2; CHPF2; SMARCD3* |
| 8:103542538-103550211 | rs13250010 | G0 | T | G | 0.356 | 0.063 | 0.01 | 7.71E-11 | 0.062 | 0.015 | 3.47E-05 | *KB-1980E6.3* | *UBR5; RRM2B; ODF1; KB-1980E6.3* |
| 9:32933492-33385427 | rs13297246 | G2 | A | G | 0.184 | 0.195 | 0.013 | 4.21E-55 | 0.202 | 0.019 | 2.36E-26 | *B4GALT1* | *B4GALT1* |
| 11:65555524-65555524 | rs10896045 | G1 | A | G | 0.301 | 0.079 | 0.013 | 2.61E-09 | 0.079 | 0.017 | 2.42E-06 | *OVOL1* | *OVOL1; AP5B1* |
| 17:16813994-16875636 | rs34562254 | G1 | A | G | 0.106 | 0.166 | 0.019 | 1.48E-18 | 0.092 | 0.024 | 9.95E-05 | *TNFRSF13B* | *TNFRSF13B* |
| 17:43856639-44863413 | rs199516 | G0 | T | C | 0.221 | 0.068 | 0.012 | 1.20E-08 | 0.058 | 0.018 | 1.25E-03 | *WNT3* | *ARHGAP27; CRHR1; SPPL2C; MAPT; KANLS1; ARL17B; LRRC37A; NSF; WNT3* |
| 17:45766846-45870129 | rs1808192 | G2 | A | G | 0.351 | 0.061 | 0.01 | 5.94E-10 | 0.051 | 0.015 | 6.78E-04 | *TBKBP1* | *TBKBP1; TBX21* |
| 17:56404349-56418136 | rs2526377 | G2 | A | G | 0.463 | 0.062 | 0.009 | 5.59E-11 | 0.057 | 0.015 | 8.05E-05 | *BZRAP1* | *BZRAP1; SUPT4H1; RAD5C1* |
| 17:79158040-79268562 | rs2659005 | G2 | T | C | 0.436 | 0.079 | 0.009 | 4.36E-17 | 0.097 | 0.016 | 6.99E-10 | *SLC38A10* | *AZI1; ENTHD2; SLC38A10; C17orf89* |
| 21:36564553-36665202 | rs4817708 | G0 | T | C | 0.229 | 0.073 | 0.011 | 2.39E-11 | 0.043 | 0.017 | 1.15E-02 | *RUNX1* | *RUNX1* |

### Discovery and replication GWAS

We performed a discovery GWAS of IgG galactosylation in seven cohorts of European descent (N=13,705). The association between HRC-imputed SNPs and three phenotypes (G0, G1 and G2) was studied under the assumption of an additive linear model. The inverse-variance meta-analysis of GWAS summary statistics resulted in 16 genome-wide significant (p ≤ 2.5 × 10^−8^) genomic loci associated with at least one galactosylation trait (Table 1). Quantile-quantile plots of associations for three traits are shown in Fig. S1.

For twelve loci, the same trait-SNP association reached the significance threshold p-value ≤ 0.0031 (P ≤ 0.05/16 loci) in replication meta-analysis (N=7,775). The association between the top SNP in the HLA region (chr6:31107733-31164511) was not replicated due to the unavailability of the SNP and SNPs in LD in all four replication cohorts. However, the association of the HLA region and IgG glycan patterns was observed in the previous GWAS of IgG glycome^27,28,30,31^. For top SNPs in three regions (chr6:74168723-74285118, chr4:103390496-103567348 and chr21:36564553-36665202) the association was not significant after correction for multiple testing in the replication meta-analysis, but the direction of the effect was consistent with the discovery analysis (Table 1).

Due to lack of compatibility in glycan trait definition in the current study and previous GWAS of IgG glycome^27–31^, we checked for the overlap of the associated genomic regions across any previously examined IgG glycan traits. Thirteen loci overlap with previously identified and replicated genomic regions (Appendix Table 2). The remaining three associations in chr4:103390496-103567348, chr17:56404349-56418136 and chr6:74168723-74285118 are considered as novel.

Among 16 loci, three loci were G2-specific (rs3822960, rs1808192 and rs2526377), two were G1-specific (rs10896045 and rs34562254) and two were G0-specific (rs13250010 and rs199516) (Fig. 1a), which is intriguing since conversion of G0 to G1, and then G2 is a consequence of the activity of the single enzyme (product of *B4GALT1* gene).

**Fig. 1:**
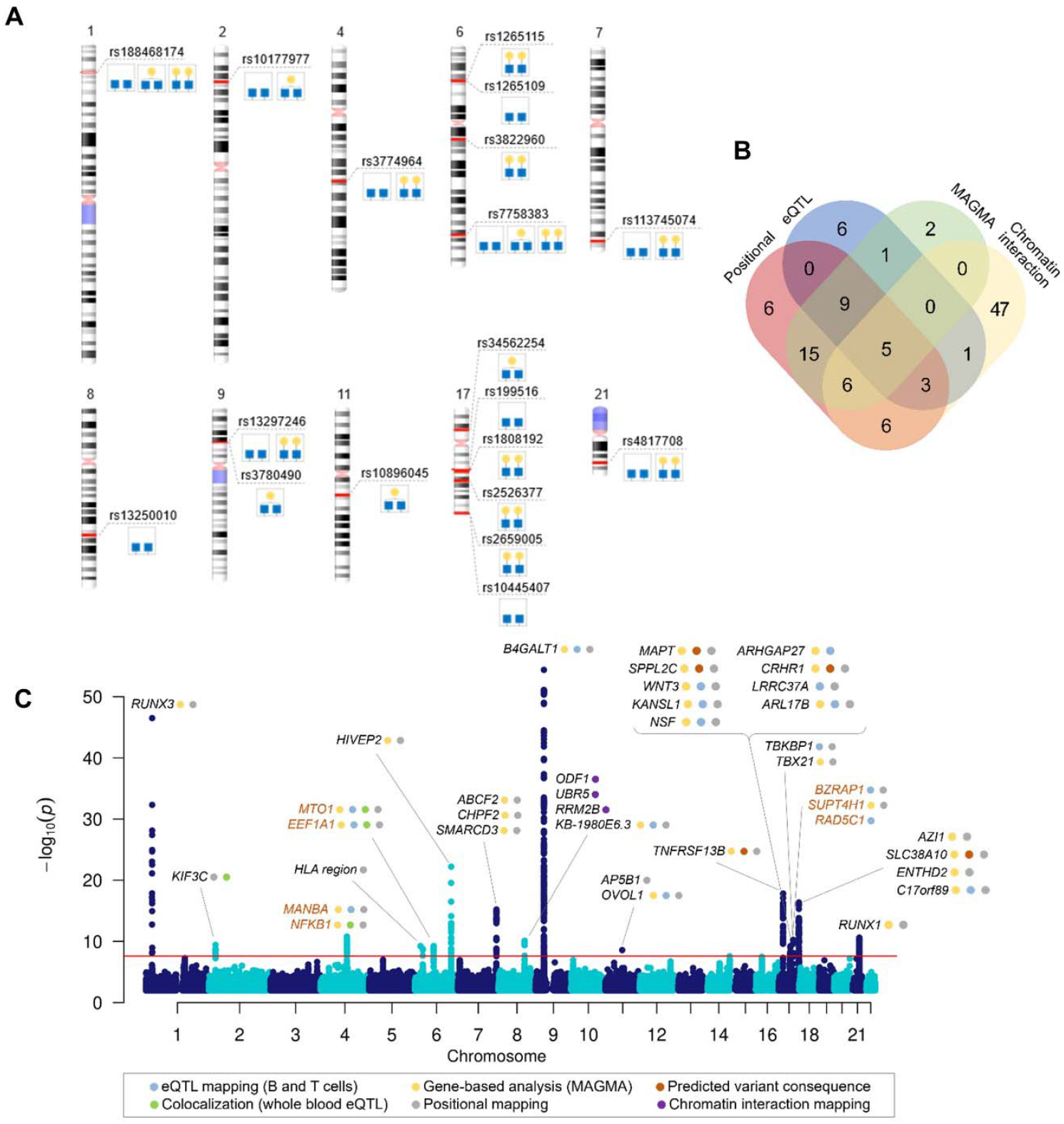
A) Top SNP in identified genomic regions for each associated trait, B) Venn diagram showing the number of genes mapped by positional mapping, chromatin interaction mapping, eQTL mapping and genome-wide gene-based association analysis (MAGMA), C) Manhattan plot of genome-wide significant associations in IgG galactosylation GWAS with prioritized genes in each locus. Plot shows -log10(p-values) of association on y-axis and SNPs ordered by chromosomal location on x-axis. Red line indicates the genome-wide significance threshold (2.5 ×10^−8^). Orange gene names indicate novel loci associated with IgG glycosylation.

Before functional evaluation of GWAS hits, careful inspection of the genomic regions was undertaken to prioritize and choose the genes with strong evidence for a potential role in a biological pathway. We applied a set of *in silico* methods to prioritise genes for the functional analysis. First, we mapped associated variants to genes using mapping strategies based on position, overlap with eQTL associations and chromatin interaction as implemented in FUMA^34^, and identified 107 candidate genes in total (Appendix Table 3). The number of genes prioritised by two or more approaches is shown in Fig. 1b.

The gene analysis in MAGMA is based on a multiple linear principal components regression model in which the statistic is computed by combining the p-values of the individual SNPs in a gene while taking into account their LD structure. In gene analysis, SNP data is aggregated to the whole gene level to test the joint association of all SNPs in the gene with the phenotype, thereby making it possible to detect effects consisting of multiple weaker SNP-phenotype associations that would otherwise be missed. The gene-based association test for G0, G1 and G2 identified 38 genes significant at FDR < 0.05 (Appendix Table 4).

Given that missense variants have a direct impact on protein function and therefore a more straightforward effect on the phenotype, we also assessed the functional consequences of associated variants using Variant Effect Predictor^35^. For two genes, *SLC38A10* and *MAPT*, variants with potentially pathogenic amino acid (aa) change were detected by both SIFT and Polyphen2 algorithms. For additional four genes, *MANBA, SPPL2C, CRHR1* and *TNFRSF13B*, the variants were predicted to result in a benign aa change. The details on position and aa changes are listed in Table S1.

Finally, to assess the potential pleiotropic effects of the variants on IgG galactosylation and gene expression we performed colocalization analysis with the whole blood eQTLgen dataset^36^. By comparing the regional association patterns with the approximate Bayesian Factor approach in coloc^37^ we identified five genes (*KIF3C, NFKB1, MIR-142, EEF1A1* and *MTO1*) whose expression patterns colocalize with IgG galactosylation (PP4 > 75%) (Table S2, Fig. S2). Previous prioritization efforts in GWAS of IgG glycosylation, as well as implications for gene’s potential function in glycosylation process were taken into account in the final gene prioritization, which resulted in 37 credible gene candidates in the identified regions (Fig. 1c). Table S1 contains details on prioritization evidence for each gene.

### IgG galactosylation and diseases

Due to existing evidence of altered IgG glycosylation patterns in multiple conditions^11^, we assessed the pleiotropy of the IgG galactosylation-associated loci with a range of autoimmune, inflammatory and infectious diseases. The threshold of 75% and higher was applied for the posterior probability for colocalization of traits (PP4). A shared association pattern between SLE and monogalactosylation (PP4 = 82%) was detected in a locus on chromosome 11 (chr11:65555524), while agalactosylation-associated locus chr17:43856639-44863413 colocalized with breast cancer (PP4 = 95%), COVID-19 hospitalization (PP4 = 93%), SLE (PP4 = 89%) and schizophrenia (PP4 = 84%) (Table S3). Because of the extensive LD pattern, there are more than twelve candidate genes in the chromosome 17 locus. It is important to note that high PP4 indicates at least one shared causal variant and low PP4 does not mean that two phenotypes do not share one, especially if the probability of both phenotypes being associated with the region but having different causal variants (PP3) is high.

### Polygenic score

A polygenic score (PGS) aggregates the effects of many genetic variants into a single number which predicts genetic predisposition for the phenotype. In the standard approach, a PGS is a linear combination of linear regression effect estimates and allele counts at single-nucleotide polymorphisms (SNPs).

For each of the G0, G1 and G2 traits, we created two PGS models based on genome-wide association meta-analysis (GWAMA) results. Specifically, we derived the first model based on the SBayesR ^38^ method, and the second model using a clumping and thresholding (C+T) method. These models were tested within the CEDAR dataset, which has aggregated genotype data and galactosylation information on 162 participants of European ancestry and which was not used in the GWAMA.

The estimated SNP-based heritability (h2SNP, SBayesR package^38^) was 36.5% for the G0, 27.2% for G1, and 36.8% for the G2 trait. Our best PGS models explained 7.4% [p-value: 4.79 × 10^−4^], 2.2% [p-value: 0.059] and 7.4% [p-value: 4.77 × 10^−4^] of G0, G1 and G2 variance in CEDAR samples. The complete results of the implementation of SBayesR and C+T models can be found in Appendix Table 5.

### Functional validation of GWAS hits

The genes for functional follow-up were selected based on novelty in GWA studies for IgG galactosylation and by reviewing the evidence for prioritization, expression in B cells (Human Protein Atlas), as well as known roles of the encoded proteins. Newly discovered GWAS hits, *EEF1A1* and *MANBA*-*NFKB1*, where *NFKB1* and *MANBA* are found in the same locus, were selected and functional follow-up was performed to decipher which one is indeed involved in galactosylation. We also included *HIVEP2* because it was previously assigned in the gene network as a transcription factor potentially involved in the regulation of *B4GALT1* gene expression^30^, and *TNFRSF13B* for its highly specific expression in B cells and function in humoral immunity. *SLC38A10* was included as one of the genes prioritized in chr17:79158040-79268562 locus to test its potential function as a transporter protein in the IgG glycosylation pathway. We also included *KIF3C* as it was the prioritized gene in chr2:26109539-26149988 locus.

To functionally validate the potential role of the selected GWAS hits in IgG galactosylation, we used the newly developed transient expression system HEK293FS for IgG production with stably integrated dSaCas9-VPR or dSpCas9-KRAB expression cassettes for target gene upregulation or downregulation^32^. Therefore, the candidate genes were manipulated in dCas9-VPR or dCas9-KRAB containing monoclonal cell lines transiently cotransfected with a plasmid carrying genes for IgG heavy and light chains and specific gRNA. Subsequently, secreted IgG was analysed for glycan phenotype.

Utilizing dCas9-VPR cell line and three gRNAs for each locus, we successfully upregulated all selected loci except *SLC38A10* and *HIVEP2* (Fig. S3a, Table S4). Successful downregulation in the dCas9-KRAB cell line was accomplished using 3 gRNAs for all selected loci except *NFKB1* and *HIVEP2* (Fig. S3b). A significant change of IgG glycome composition was observed after dCas9-VPR targeting of *HIVEP2, MANBA, TNFRSF13B* and *EEF1A1* (Fig. 2, Appendix Table 6). Interestingly, manipulation of the *HIVEP2* promotor did not upregulate *HIVEP2* transcript level sufficiently to reach statistical significance at transcript level, but it resulted in a significant increase of digalactosylated structures and a concomitant decrease of agalactosylated structures on IgG (Fig. 2a). Upregulation of the *MANBA* locus resulted in a significant decrease of monogalactosylated IgG glycans, but no change in levels of agalactosylated and digalactosylated glycans was observed (Fig. 2b). Targeting *TNFRSF13B* in dCas9-VPR cell line resulted in almost 400-fold increase of its expression and a moderate increase in agalactosylated IgG glycans (Fig. 2c). Upregulation of the *EEF1A1* locus resulted in a significant decrease of monogalactosylated glycans and in a concomitant increase of agalactosylated IgG glycans (Fig. 2d).

**Fig. 2.**
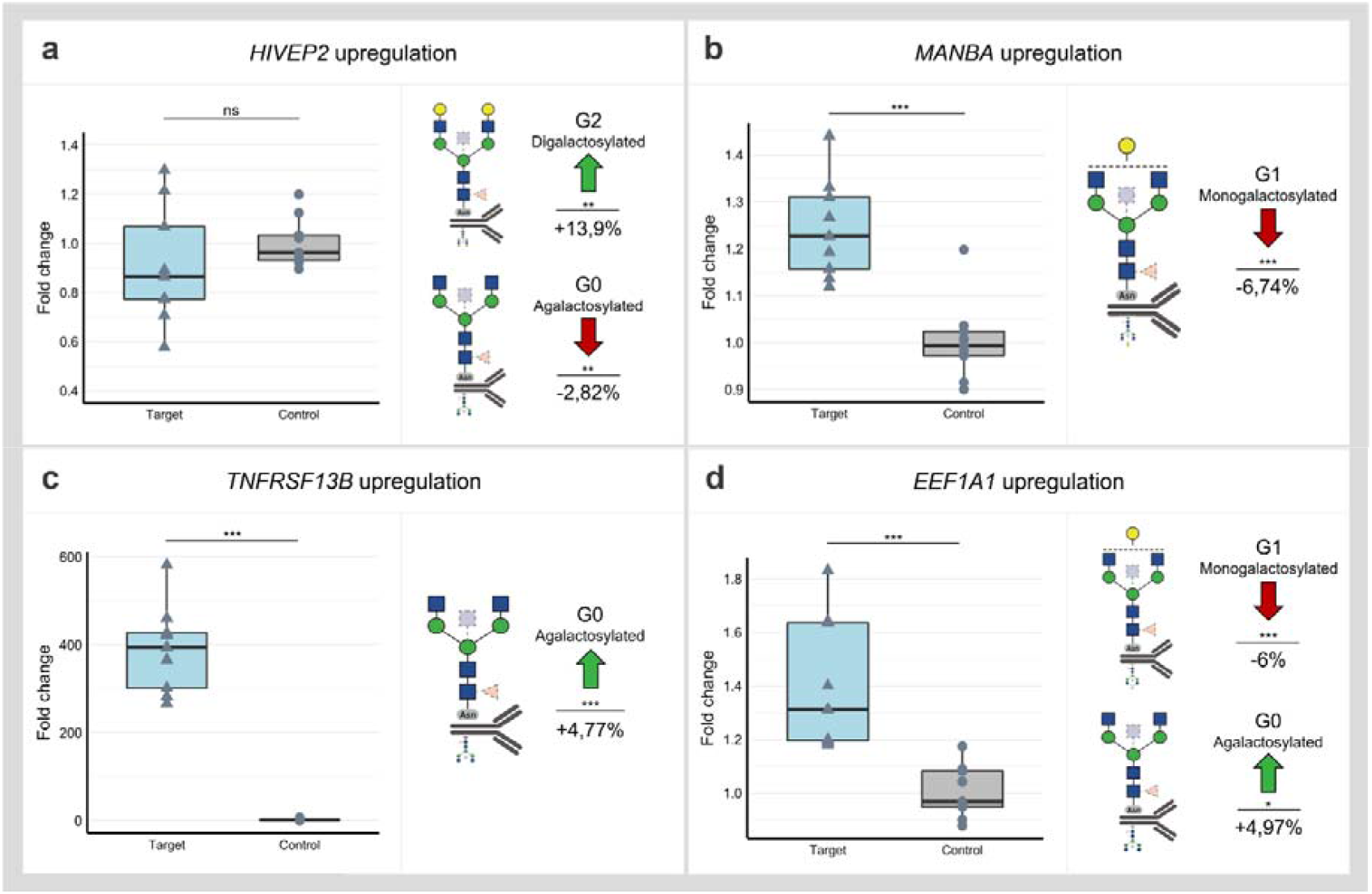
Targeting of selected GWAS loci associated with IgG galactosylation (*HIVEP2, MANBA, TNFRSF13B* and *EEF1A1)* in dCas9-VPR monoclonal cell lines resulted in significant changes in IgG glycan composition. Samples containing non-targeting gRNAs served as controls. Changes in transcript levels are given as fold change values and changes in IgG phenotype are given as a relative change compared to control samples. **(a)** Manipulation of the *HIVEP2* gene did not result in a statistically significant change in *HIVEP2* transcript level, however, did induce a significant increase of digalactosylated structures (G2) with a concomitant decrease of agalactosylated IgG glycan structures (G0). **(b)** Targeting of *MANBA* by dCas9-VPR elevated transcription level of this gene which resulted in decrease of monogalactosylated IgG glycan structures (G1) **(c)** Targeting *TNFRSF13B* by dCas9-VPR resulted in ∼ 400-fold increase of transcript levels and significant change of IgG agalactosylated IgG glycans (G0). **(d)** Successful upregulation of the *EEF1A1* locus was followed by an increase of agalactosylated IgG glycans (G0) with a concomitant decrease of monogalactosylated IgG glycans (G1). Nominal p-value: *<0.05; **<0.01; ***<0.001; ns, not significant.

## Discussion

Using GWAS approach we identified 16 genomic regions associated with IgG galactosylation, which is the molecular basis of the glycan clock of ageing. Thirteen out of 16 loci were associated with traits related to IgG glycosylation in our previous GWAS studies, while three were novel^27–31^. Contrary to previous studies, in which we were looking for genes that were associated with any aspect of IgG glycosylation, in this study we focused on galactosylation, a functionally well-defined element of IgG glycosylation that is the basis of increased inflammation in people with accelerated glycan ageing. This was done by converting percentages of individual glycans in the glycome into sums of glycan structures containing either one, two or no galactose residues. This enabled us to combine GWA studies for which the phenotype analysis was performed using different analytical methods. We also estimated the SNP-based heritability to be 36.5%, 27.2% and 36.8%, for G0, G1 and G2, respectively, an estimate that very closely replicates recent heritability estimate for biological age estimate based on these glycan structures^39^. Upon prioritization efforts which resulted in 37 candidate genes, we evaluated functional relevance of these genes using HEK293FS transient expression system for IgG production with stably integrated dCas9-KRAB or dCas9-VPR fusion. Results of this study expanded the list of genes for which the functional relevance for *in vitro* expression and glycosylation of IgG was confirmed with *MANBA, TNFRSF13B* and *EEF1A1* (Fig. 2). Thus, out of 13 replicated GWAS hits, for six of them we have functionally confirmed that they act at IgG glycosylation in a way that is conserved between plasma cells and our artificial *in vitro* expression system in HEK293FS cells (Fig. 3). It is likely that some additional GWAS hits may be plasma cell specific and will be confirmed once an efficient system for gene manipulation and IgG production in B cells is developed. However, it is important to note that IgG glycosylation can be affected at multiple other levels (e.g. B-cell selection, maturation, expansion, etc.), thus the absence of functional confirmation of other loci in this assay does not mean that they are not implicated in IgG glycosylation in some other, more complex, manner.

**Fig. 3.**
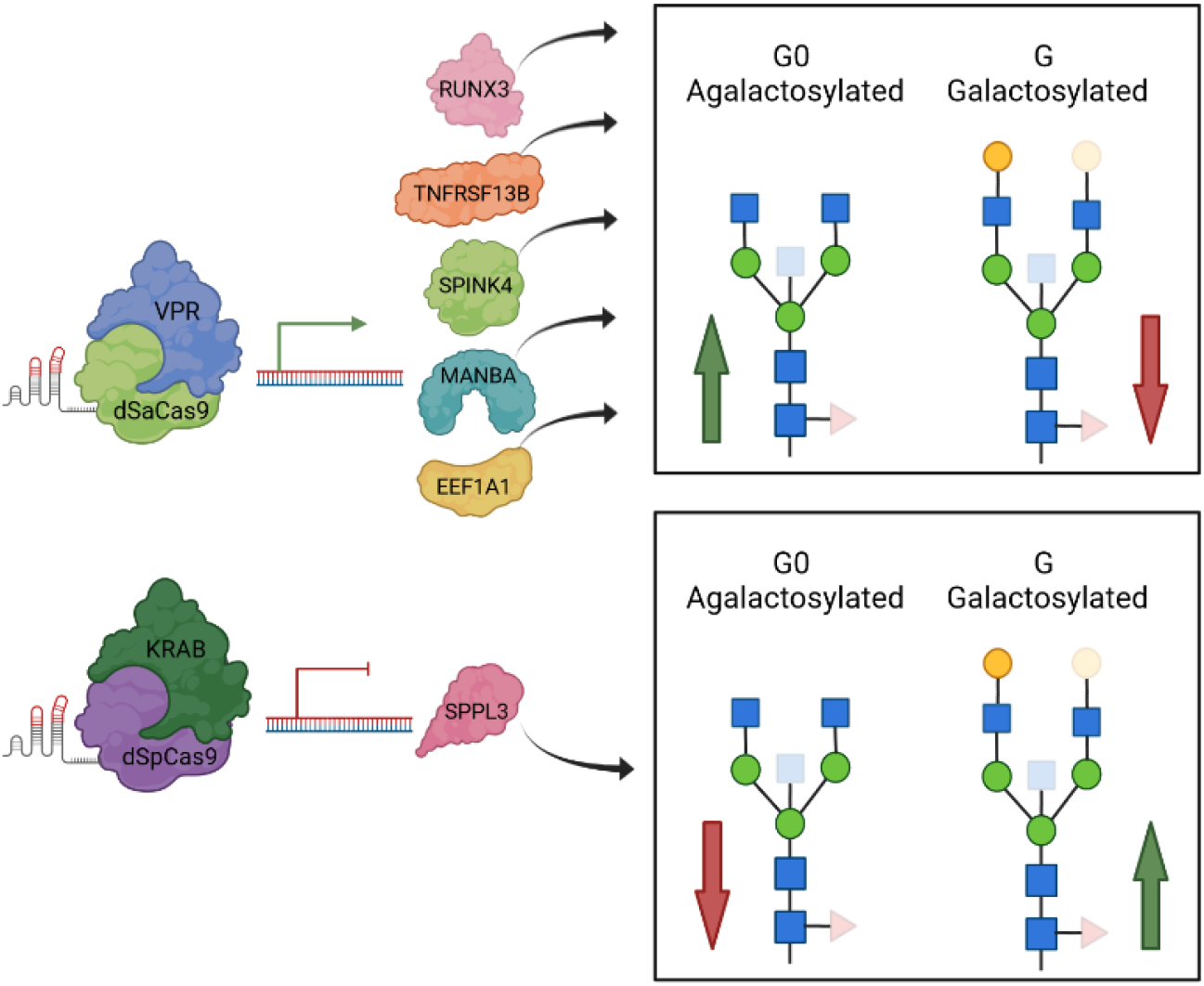
A graphical review of up-to-date functional validation of GWAS hits associated with IgG glycosylation using HEK293FS transient expression system with stably integrated dCas9-VPR or dCas9-KRAB fusions. When specific gRNA targeted dCas9-VPR to two estrogen associated genes, *RUNX3* and *SPINK4* (Mijakovac et al. 2021), as well as to the *TNFRSF13B, MANBA* and *EEF1A1* loci, transcription was upregulated that resulted in decreased levels of galactosylated (red arrow within box), increased levels of agalactosylated IgG glycan structures (green arrow within box) or both. Downregulated transcription of *SPPL3* resulted in an increase in galactosylated structures with a concomitant decrease in agalactosylated IgG glycan structures. Protein structures do not depict true protein structures in humans, generic protein shapes are chosen for easier visualization. Figure was created with BioRender.com. Accessed on 16 November 2021.

The total heritability of G0, G1, G2 is not known, however, the heritability of individual IgG glycan peaks ranges between 27% and 76%^26^. More than half of the studied glycan traits have additive genetic component estimate of 50% which would imply that half of the variance in their values could be explained by common variants, an observation similar to other complex traits (Menni et al., 2013). The constructed PGS models explained 7.4% of the variance in G0 and G2 traits in an out-of-sample testing, thus capturing about 20% of the estimated SNP heritability. However, for G1 the PGS model explained a minor proportion of SNP heritability. The explanation why for G1, that had comparable SNP heritability, the proportion of variance explained was much less may lie in a flatter distribution and hence individually smaller genetic effects onto G1; and observation which may be described as “higher polygenicity”. Indeed, while for G0 and G2 lead associations explained about 1.5% of variance, for G1 the lead variant near *TNFRSF13B* explained only about 0.5% of its variation. With smaller individual effects, one needs larger discovery GWAS to construct powerful PGS.

Given that the majority of identified candidate SNPs are located in non-coding regions, we applied several *in silico* approaches to prioritize potential causal genes in associated genomic regions, including exploring functional consequences of associated variants, pleiotropy with gene expression, gene-based association analysis, as well as evidence from previous GWA studies and prior biological knowledge of gene function. Among 37 candidate genes from 16 associated loci, for the two genes, *RUNX3* and *B4GALT1*, the effect on IgG glycosylation has recently been demonstrated using direct gene expression modulation in HEK293FS IgG transient expression system^32,40^.

Due to existing evidence of altered IgG glycosylation patterns in multiple conditions^11^, we performed colocalization analysis with a range of diseases and observed a high probability for pleiotropic effects in the *AP5B1/OVOL1* locus on chromosome 11 for SLE and monogalactosylation. Interestingly, for the same locus, we observed considerable probability for colocalization (PP4=0.56) with asthma and allergy. Although this is below our pre-defined posterior probability for colocalization (PP4) threshold of PP4 > 0.75, the finding is in line with previous results linking this locus with variation of specific digalactosylated structures (FA2FG2S1), expression of the *OVOL1* gene and the risk of asthma^31^. *OVOL1* gene encodes a putative zinc finger containing transcription factor with a role in the development and differentiation of epithelial and germ cells^41^. The locus on chromosome 17 was pleiotropic for agalactosylation and SLE, breast cancer, COVID-19 hospitalization and schizophrenia. The colocalized region on chromosome 17 (chr17:43856639-44863413) contains at least twelve genes and is known for numerous copy number variants. It also includes a megabase-long inversion polymorphisms (H1 and inverted H2 forms) which contain partial duplication of *KANSL1* gene with inverted H2 isoform having a higher frequency than H1 isoform in the European population^42^. The complexity of the locus makes it challenging to prioritise a specific gene without further investigation.

The functional follow-up was based on the previously developed HEK293FS transient expression system for IgG production which was shown to be an excellent tool for studying the role of the genes identified in glycosylation GWAS^32^. Despite the fact that we could not demonstrate upregulation of *HIVEP2* mRNA using dCas9-VPR, a significant increase in digalactosylated structures with a concomitant decrease in agalactosylated structures was measured from IgG secreted from the same cells. *HIVEP2* (*HIVEP zinc protein 2*) encodes a transcription factor that interacts with numerous viral and cellular promoters, and has an established role in immunity^43,44^. Even though we prioritised *HIVEP2*, the locus interacts with variants in six genes via chromatin interactions (Fig. S4), including *FUCA2* (alpha-L-fucosidase) which is located 0.6 megabases away. It is plausible that dCas9-VPR targeting *HIVEP2* promoter disrupted these chromatin interactions which in turn altered the expression of other genes mapping to this region. In the case of the locus on chromosome 4, the functional study was utilized as one of the steps to decipher which of the two genes might influence galactosylation process as both *NFKB1* and *MANBA* were prioritised with *in silico* approaches. The *MANBA/NFKB1* region was identified in previous studies as a disease susceptibility locus for primary biliary cholangitis (PBC)^45^ and a recent study found a decreased level of IgG galactosylation in PBC patients^46^. The *NFKB1* gene encodes a subunit of the nuclear factor of kappa light polypeptide gene enhancer in B-cells (NF-κB) transcription factor family, known for its critical role in cell survival and inflammation^47^. The upregulation of *NFKB1* using dCas9-VPR resulted in a significant change in transcript level, but no change in IgG glycoprofile was observed. On the other hand, the upregulation of the *MANBA* gene expression resulted in a decrease of IgG monogalactosylated species. The *MANBA* gene encodes beta mannosidase which is an exoglycosidase enzyme cleaving beta-mannose residues from the non-reducing end of N-linked glycans^48^. As such, it has a known role in glycosylation process but currently we cannot speculate about the exact mechanism by which it affects IgG galactosylation. Interestingly, mutations in *MANBA* are known to affect kidney function^49^, blood pressure^50^ and cardiovascular disease^51^, while the monogalactosylated IgG glycan (G1) (that *MANBA* affects in our *in vitro* expression system) was previously identified as the best predictor of future cardio vascular events in women^52,53^. While at the moment do not know putative mechanisms that could explain the link between *MANBA*, G0 and cardiovascular or renal diseases, these intriguing associations warrant future studies.

The *EEF1A1* (*Eukaryotic Translation Elongation Factor 1 Alpha 1*) gene encodes for an isomer of an alpha subunit of the eukaryotic translation elongation factor-1 complex that has a role in binding of aminoacyl tRNA to the A site on ribosomes during protein synthesis ^54^. Together with EEF1A2, it is the second most abundant protein (1%-3%) in the cell with other roles outside of protein synthesis, including protein degradation, apoptosis modulation, oncogenesis and viral pathogenesis^55,56^. Previous study has shown that EEF1A1 also serves as a signal transducer during inflammation, where it can enhance interleukin 6 expression through STAT3 and PKCδ^57^. Interleukin 6 is implicated in different autoimmune and inflammatory diseases^58^ and its high levels lead to decreased IgG galactosylation^59^. By targeting dCas9-VPR to *EEF1A1*, we increased its expression level, which resulted in a decrease of galactosylated IgG glycans. Therefore, our results are concordant with the previous findings, suggesting that *EEF1A1* upregulation could lead to pro-inflammatory modulation of IgG by decreasing its galactosylation.

A locus harbouring *TNF receptor superfamily member 13B* (*TNFRSF13B*) gene was previously identified in the multivariate IgG glycosylation GWAS^31^, while here it was associated with IgG monogalactosylation. *TNFRSF13B* encodes a transmembrane activator calcium modulator and cyclophilin ligand interactor (TACI) protein, a lymphocyte-specific member of the tumour necrosis factor (TNF) receptor superfamily, which has a role in signalling pathway leading to B cell differentiation and antibody production^60,61^. Common genetic variation in *TNFRSF13B* locus was previously associated with different levels of immunoglobulins in serum^62^, while rare deleterious variants were implicated in common variable immunodeficiencies and IgA deficiencies^63,64^. Therefore, we speculate that *TNFRSF13B* might impact glycosylation profile of the sample by controlling secretion of certain IgG glycoforms. Despite extensive increase in *TNFRSF13B* expression upon activation with dCas9-VPR, we observed only a moderate increase in IgG agalactosylation and no changes in mono-or digalactosylation.

Our experimental setting might be limited by the B-cell specific function of *TNFRSF13B* and thus, we might not be able to observe changes comparable to changes in the gene expression level.

The role of four other prioritized genes, *NFKB1, KIF3C, HIVEP2* and *SLC38A10*, could not be validated in our HEK293FS transient expression system. Even though this cell system represents a good model for studies of IgG glycosylation regulation, it has some limitations. HEK293FS cells do not inherently secrete immunoglobulins and their epigenetic context is not equivalent to plasma and B cells. In addition, alternative galactosylation might depend on gene expression in other cell types and their interactions, hence we might not observe the natural effects after manipulation of gene expression in this model system, due to the lack of the right biological context.

In conclusion, in the present study we mapped 16 genetic loci regulating glycan structures that are not only the basis of the glycan clock of ageing, but also functional effectors of inflammation at multiple levels. Furthermore, we were able to confirm the functional role of three genes (*MANBA, TNFRSF13B* and *EEF1A1*) in the IgG galactosylation pathway by use of the HEK293FS transient expression system with stably integrated dCas9 fusion designed for this purpose. Fig. 3 summarizes all GWAS hits with previously unknown link to IgG glycosylation, whose functional relevance has been shown using this cell system. Further research is needed to fully elucidate functional mechanism behind their role in ageing and to reveal the complete network of gene interactions regulating the complex process of IgG glycosylation. However, this study is an important step in this direction since it clearly indicates that regulation of IgG galactosylation extends far beyond the expression of the galactosyltransferase gene that adds galactose.

## Materials and Methods

The participating cohorts were approved by local research ethics committees and informed consent from the participants was obtained. Information about sample numbers, sex and age for each cohort is provided in Appendix Table 7. Further information on genotyping, genotype imputation and quality control can be found in Appendix Table 8.

### Glycan measurements

An overview of cohorts and technologies used for IgG glycome measurement (LC-MS or UPLC), as well as the published studies where IgG glycome analysis for the corresponding cohort was described in more detail, are shown in Appendix Table 7.

Briefly, for UPLC-based analysis, IgG was isolated from blood plasma samples using protein G plates, followed by neutralization and denaturation of the protein. N-glycans were enzymatically released from IgG and fluorescently labelled with 2-aminobenzamide dye, followed by separation and quantification by hydrophilic interaction UPLC which resulted in 24 peaks (GP1-GP24), most of which represent a single glycan structure. In cases where multiple glycan structures are found under one peak, the most abundant structure is considered. The list of all glycans and their proportion under each peak can be found in Pučić et al.^65^.

For LC-MS-based glycan analysis, IgG was isolated from blood plasma samples by affinity chromatography binding to protein G plates and treated with trypsin to allow cleavage of IgG at specific amino acid sites. The cleavage resulted in different glycopeptides across IgG subclasses, thereby enabling subclass-specific glycan measurements. IgG subclass separation and detection were performed using a nano-LC system coupled with quadrupole-TOF-MS. IgG2 and IgG3 subclasses have the same tryptic glycopeptide moieties, thus the separation of the subclass-specific glycopeptides was not possible. LC-MS quantification resulted in 50 values which correspond to 20 glycans measured on IgG1, 20 glycans on IgG2/3 and 10 glycans on IgG4. The list of glycan structures measured by UPLC and LC-MS along with their description is listed in Appendix Table 9 and 10.

### Glycan data pre-processing

Prior to genetic analysis, the best approach for harmonization of glycan measurements by UPLC and LC-MS was chosen (Supplementary Methods) to allow for meta-analysis of GWAS summary statistics from all cohorts. To reduce the impact of experimental variation on the downstream analysis, glycan measurements were normalized using median quotient normalization and batch corrected using empirical Bayes method^66^ implemented in ComBat function of the “sva” package^67^ in statistical software R^68^. Median quotient normalization was applied due to negligible differences between different normalization approaches in the comparison and further supported by previous recommendation^33^. Next, three derived traits were calculated as the percentage of structures in the total IgG glycome containing zero, one or two galactose units as G0, G1 and G2, respectively (Appendix Table 11). In this way, the subclass-specific glycan values from LC-MS measurements were summarized to allow for comparison with total glycan data obtained with UPLC. The glycan data was pre-processed centrally in Genos for all cohorts except Leiden Longevity Study (LLS), for which the glycan data was pre-processed by the phenotype provider from the Leiden University Medical Center.

### Genome-wide association study

Discovery genome-wide association study was performed in seven cohorts of European descent (CROATIA-Vis, CROATIA-Korcula, CROATIA-Split, TwinsUK, EPIC, VIKING, ORCADES) with a total sample size of 13,705. Quantile normalization was applied to the derived traits in each cohort. For CROATIA, VIKING and ORCADES cohorts, first linear mixed model was fit to adjust for age, sex and cohort-specific covariates while also accounting for the genomic kinship. This step was performed using polygenic function in GenABEL R package^69^, followed by testing the association between SNPs and phenotype using the RegScan software v0.5^70^. For TwinsUK cohort, the derived traits were adjusted for age and sex and the association analysis was carried out using GEMMA^71^ including kinship matrix to correct for family structure. SNPTEST software was used for association testing of age- and sex-adjusted glycan values with genetic variants in EPIC cohort. GWAS was performed on Haplotype Reference Consortium (HRC)^72^ imputed genotypes, assuming an additive linear model of association. Quality control of study-specific summary statistics was performed prior to meta-analysis using EasyQC R package as described in Winkler et al.^73^

After quality control, the GWAS summary statistics for seven cohorts were pooled using the inverse-variance weighted method for meta-analysis as implemented in METAL software^74^. METAL software also allowed for the estimation of genomic control (GC) to correct test statistics to account for population stratification in the studies included in the meta-analysis (lambda GC median =1.00; range 0.98-1.01). Given that we performed GWAS for three correlated traits, we used 5 × 10^−8^/2= 2.5 × 10^−8^ as a genome-wide significance threshold, where 2 denotes the number of PCs that explain 99% of the variation in the three galactosylation traits.

### Replication analysis

Following the discovery GWAS, we performed replication meta-analysis to validate results from the discovery phase. Four independent cohorts of European descent were used: KORA F4, LLS, EGCUT and GCKD, with a total sample size of 7,775. Glycan measurements for KORA F4 and LLS cohort were quantified with LC-MS, while EGCUT and GCKD glycan measurements were obtained with UPLC. The strongest glycan trait-SNP pair from each locus in the discovery analysis was meta-analysed using the fixed-effect inverse variance method. The significance threshold was set to p-value < 0.05/16 = 0.0031, where 16 is the number of associated genomic regions. We also checked the consistency of effect direction between the discovery and replication analyses in cases where the top SNP did not formally replicate.

### Genomic loci definition

Genomic loci associated with galactosylation phenotypes were defined using FUMA v. 1.3.6b^34^. SNP2GENE function in FUMA first identifies independent (LD r^2^ < 0.6) significant SNPs with MAF > 0.01 to determine borders of genomic locus. LD estimates were inferred from reference panel derived from 10,000 subjects of European descent in UK Biobank (Bycroft et al., 2018). The maximum distance of 250kb was used for merging LD blocks into a single genomic locus. SNPs in LD (r^2^ > 0.6) with independent SNPs within 250 kb distance were selected as associated SNPs and considered in further analysis.

### Gene prioritization

Since we aimed to demonstrate *in vitro* the functional activity of the proteins encoded by genes potentially causal for IgG galactosylation, we first employed multiple *in silico* strategies to prioritise candidate genes. Briefly, we used the following criteria: gene mapping (prioritising based on genomic position, overlap with eQTLs and chromatin interactions), functional consequence-based prioritisation (genes whose missense variants were associated with galactosylation), colocalization of SNP associations with gene expression in whole blood and with galactosylation, and gene-based analysis.

### Gene mapping

Genes in the significantly associated genomic regions were mapped using three approaches implemented in FUMA: positional mapping, eQTL mapping and chromatin interaction mapping. Genes were positionally mapped based on ANNOVAR^76^ annotations and the maximum distance of <10 kb between associated variants and genes. The eQTL mapping was based on overlap of galactosylation-associated variants and eQTLs in B and T cells, including Database of Immune Cell Expression (DICE)^77^, Fairfax *et al*.^78^ and CEDAR^79^ datasets. Only significant eQTL signals at FDR < 0.05 were considered. Chromatin interaction mapping was performed using the Hi-C data derived from B cell line (GM12878) ^80^ and the suggested FDR value < 1 × 10^−6^ for significant chromatin interaction was used.

### Functional consequences

To assess potential functional consequences of galactosylation associated variants we used SIFT and Polyphen-2 algorithms, as implemented in the Variant Effect Predictor (VEP) v97 by Ensembl^35^.

### Colocalization with gene expression in whole blood

The colocalization of galactosylation GWAS signals and gene expression was estimated using Approximate Bayes Factor (ABF) method^81^, as implemented in the coloc package^37^ in R. The method outputs five posterior probabilities, one for each of the five hypotheses: H0) no association with either of the two traits, H1) association with trait 1, but not with trait 2, H2) association with trait 2, but not with trait 1, H3) association with both trait 1 and trait 2, but two independent causal variants and H4) association with both trait 1 and trait 2 and one shared causal variant. Whole blood eQTL data was obtained from the publicly available eQTLgen dataset which was derived from 31,684 individuals across 37 cohorts^36^. Tissue- or cell-specific eQTL data (e.g. B cell or T cell) were not used due to low number of SNPs in the associated regions which overlap in galactosylation GWAS summary statistics and tissue-specific eQTL data to allow for reliable colocalization test. The posterior probabilities for colocalization with cis-eQTLs were computed for each gene found in galactosylation-associated genomic loci using SNP p-values and MAF. The default values for prior probabilities were used (p1D□=□1 × 10^−4^, p2□=□1 × 10^−4^ and p12□=□1 × 10^−5^). The threshold of 75% for PP4 (probability of the same shared variant for two traits) indicated positive colocalization and strong support for prioritization of the gene in the given genomic locus.

### Gene-based association test

Genome-wide gene-based association test for all three traits was performed by MAGMA v1.08^82^ to evaluate the joint effects of variants in 18,655 protein-coding genes while accounting for LD between those variants. In such test, SNP data is aggregated to the whole gene level to test the joint association of SNPs in the gene with the phenotype to allow detection of effects comprising of multiple weaker SNP-phenotype associations that would potentially be missed. MAGMA uses p-values derived from variant-based analyses as input and implements the SNP-wise mean model which derives a mean χ^2^ statistic for SNPs in the gene and a p-value by using a known approximation of sampling distribution. Only genes significant at FDR < 0.05 were considered.

### Colocalization with diseases and traits

The ABF colocalization method was used to explore the pleiotropy between IgG galactosylation and complex diseases for which there is previous evidence of aberrant IgG glycosylation^11^ and traits with shared associated variants in the GWAS Catalog, as obtained from GWAS Catalog query (accessed in June 2021). The full list of traits and links for summary statistics download is available in Table S5. The default values for prior probabilities were used (p1□=□1 × 10^−4^, p2□=□1 × 10^−4^ and p12□=□1 × 10^−5^). The posterior probabilities by ABF were computed using beta and standard error values if available in the dataset, otherwise, SNP p-values and MAF were used. The threshold of 75% for PP4 (probability of the shared underlying causal variant for two traits) was used for colocalization and evidence of high confidence for pleiotropy between IgG galactosylation and disease or trait.

### Polygenic score

SbayesR method reweights the effect of each variant according to the marginal estimate of its effect size, statistical strength of association, the degree of correlation between the variant and other variants nearby, and tuning parameters. This method requires a compatible LD matrix file computed using individual-level data from a reference population. For these analyses, we used publicly available shrunk sparse GCTB LD matrix including 2.8 million pruned common variants from the full UK Biobank (UKB) European ancestry (n□≈□450,000) data set and computed from a random set of 50,000 individuals of European ancestry from the UKB data set^38,83^. SbayesR was run for each chromosome separately, and with the default parameters except for the number of iterations (N=3000) and p-value (0.9) (Appendix Table 12).

Clumping and thresholding model was built using a p-value and linkage disequilibrium-driven clumping procedure in PLINK version 1.90b (--clump) ^84^. In brief, the algorithm forms clumps around SNPs with association p-values less than a provided threshold (p-value=5 × 10^−8^). Each clump contains all SNPs within 250 kilobases of the index SNP that are also in linkage disequilibrium with the index SNP as determined by a provided pairwise correlation (r^2^=0.2) threshold in the linkage disequilibrium reference. The algorithm iteratively cycles through all index SNPs, beginning with the smallest p-value, only allowing each SNP to appear in one clump. The final output should contain the most significantly disease-associated SNP for each linkage disequilibrium-based clump across the genome.

The prediction accuracy was defined as the proportion of the variance of a phenotype that is explained by PGS values (R^2^). To calculate PGS based on the PGS model we used PLINK2 software, where PGS values were calculated as a weighted sum of allele counts. Out-of-sample prediction accuracy was evaluated using samples from the CEDAR cohort that was not used for discovery or replication.

### Functional validation of GWAS hits associated with galactosylation trait

#### Plasmid constructs for targeted upregulation/downregulation of prioritized GWAS hits

Seven genes strongly associated with IgG glycosylation were selected for the follow-up functional validation. A newly developed transient expression system with stably integrated CRISPR/dCas9 fusions dSaCas9-VPR (for gene expression upregulation) and dSpCas9-KRAB (for gene expression downregulation)^32^ was used for *in vitro* validation. Three guide RNAs (gRNAs) were designed using the CRISPick online tool (Broad Institute) for adequate dCas9 orthologue (dSaCas9 or dSpCas9) and each selected locus (*KIF3C, NFKB1, MANBA, SLC38A10, TNFRSF13B, EEF1A1* and *HIVEP2*). Specific gRNAs were assembled with genes coding for IgG light and heavy chains, CBh promoter and bGH terminator using Golden Gate enzymes in three steps as described in Mijakovac *et al*.^32^. Non-targeting gRNAs were assembled in the same manner. Plasmids pORF-hp21, pORF-hp27 (Invivogen, San Diego, CA, USA) and p3SVLT were used for enhanced IgG production^85^. Specific gRNA sequences targeting each gene are shown in Appendix Table 13.

### Transfection of monoclonal dCas9-VPR/dCas9-KRAB HEK293FS cell lines

The main approach was to use suspension cell lines with integrated expression control machinery based on dCas9 fusions with IgG-producing plasmid bearing gRNA expression cassettes, as described in Mijakovac *et al*.^32^. Briefly, suspension-adapted monoclonal cell lines with stably integrated dSaCas9-VPR (dCas9-VPR) or dSpCas9-KRAB (dCas9-KRAB) were grown until they reached the appropriate density for cell transfection with gRNA-IgG bearing plasmids as well as plasmids used for enhanced protein production in mass ratio gRNA and IgG chain bearing plasmid, p3SVLT, pORF-hp21 (Invivogen, San Diego, CA, USA) and pORF-hp27 (Invivogen, San Diego, CA, USA) 0.69/0.01/0.05/0.25. Transfections were performed with 293fectin transfection reagent diluted in Opti-MEM I Reduced Serum Medium (Thermo Fisher Scientific, Waltham, MA, USA). Five days following transfection, cells were centrifugated and used for gene expression analysis, while supernatant was used for IgG isolation and glycan analysis.

### Reverse transcription and quantitative Real-Time PCR (qPCR)

Total RNA was isolated from cell pellets with Rneasy Mini Kit (Qiagen, Hilden, Germany) and 50 ng of RNA was converted to cDNA using the PrimeScript Rtase (TaKaRa, Kusatsu, Japan) and random hexamer primers (Invitrogen, Waltham, MA, USA). All samples were treated with Turbo Dnase (Invitrogen, Waltham, MA, USA) to remove any remaining DNA. Gene transcripts were detected with SYBR Green Gene Expression Assay using the 7500 Fast Real-Time PCR System. Primer sequences are listed in Appendix Table 14. The mean value of 9 replicates was normalized to *HPRT1* expression which was used as an endogenous control. Fold changes in transcript expression compared to non-targeting control were analysed using the ΔΔCt method^86^.

### Glycan measurements

IgG glycan measurements were done as described previously^32^ with a few modifications described below. Briefly, this method consisted of five main steps: 1) IgG isolation from HEK293FS transient expression system cell culture supernatants using Protein G Agarose fast flow beads (Merck, Darmstadt, Germany); 2) Enzymatic release of N-glycans from isolated IgG using PNGase F (Promega, Madison, WI, USA); 3) fluorescent labelling of released IgG glycans with procainamide hydrochloride (Thermo Fisher Scientific, Waltham, MA, USA); 4) clean-up of labelled IgG glycans using hydrophilic interaction liquid chromatography solid-phase extraction (HILIC-SPE) on a 0.2 μm Supor filter plate (Pall Corporation, Port Washington, NY, USA) and 5) separation of fluorescently labelled glycans by hydrophilic interaction chromatography on a Waters Acquity UPLC instrument (Waters, Milford, MA, USA). The IgG isolation step was modified from that previously described in the way that all washing steps after incubation of samples with Protein G beads as well as elution of IgG were done on an Orochem filter plate (Orochem Technologies Inc., Naperville, IL, USA). The obtained UPLC chromatograms were all separated in the same manner into 24 peaks and the amount of glycans in each peak was expressed as a percentage of total integrated area.

### Statistical analysis

Statistical analysis of RT-QPCR and IgG glycan data was performed using R^68^ and GraphPad (GraphPad Software, San Diego, CA, USA). RT-QPCR was done in two technical replicates and biological replicates were pooled from independent experiments. Group differences were determined using a two-tailed t-test on ΔΔCt values. IgG glycans were measured as a percentage of area under chromatogram peaks (Table S6). Derived glycan traits (G0, G1 and G2) were calculated as a sum of glycan peaks containing zero, one or two galactose units. Initial round of experiments involved both up-and down-regulation of the genes and was used as pilot study to determine whether the expression of genes changed and if there were significant changes in glycan profile by applying Student’s T-test. Five genes (*HIVEP2, MANBA, EEF1A1, TNFRSF13B* and *NFKB1*) were selected and the experiments for gene expression upregulation were repeated. To combine results of testing differences in control and treated samples by Student’s T-test across multiple experimental runs, meta-analysis approach was applied using metafor R package^87^.

## Data Availability

The full summary statistics for discovery meta-analysis for G0, G1 and G2 traits is deposited at Zenodo (10.5281/zenodo.7227999). The summary statistics for gene expression levels in different tissues and cell types are available in the eQTLcatalogue project, DICE project and eQTLGen consortium. The summary statistics for other complex traits are available in publicly available resources, as listed in Table S5. Additional data related to this paper may be requested from the authors.

https://zenodo.org/record/7227999#.ZEaBH3ZBxD8

## Acknowledgments

This work was supported by European Structural and Investment Funds grant for the Croatian National Centre of Competence in Molecular Diagnostics grant KK.01.2.2.03.0006, Croatian National Centre of Research Excellence in Personalized Healthcare grant KK.01.1.1.01.0010, IRI “CardioMetabolic” grant KK.01.2.1.02.0321, ERC-Synergy grant “GlycanSwitch” from the European Research Council grant 101071386. AFH was supported by H2020-MSCA-ITN IMforFUTURE grant 721815. The work of AN was supported by the Ministry of Education and Science of the Russian Federation via the state assignment of the Novosibirsk State University (project “Graduates 2020”). The work of SZS and YSA was partially supported by the Research Program at the MSU Institute for Artificial Intelligence. The work of LK was supported by an RCUK Innovation Fellowship from the National Productivity Investment Fund (MR/R026408/1). The work of YL was supported by the German Research Foundation (grant KO_3598/4-2 to AK). The work of AK was supported by the German Research Foundation (grant KO_3598/5-1). RRCC, CW and MBS were supported by German Ministry of Education and Research (BMBF) and the State of Brandenburg DZD grants 82DZD00302 and 82DZD03D03. CW was also supported by SciLifeLab & Wallenberg Data Driven Life Science Program grant KAW 2020.0239. TwinsUK study was funded by Wellcome Trust grant 212904/Z/18/Z, Medical Research Council AIMHY grant MR/M016560/1, European Union H2020 grant 733100. TwinsUK and MM were also supported National Institute for Health Research (NIHR)-funded BioResource, Clinical Research Facility and Biomedical Research Centre based at Guy’s and St Thomas’ NHS Foundation Trust in partnership with King’s College London.

## Data and materials availability

The full summary statistics for discovery meta-analysis for G0, G1 and G2 traits is deposited at Zenodo (10.5281/zenodo.7227999). The summary statistics for gene expression levels in different tissues and cell types are available in the eQTLcatalogue project, DICE project and eQTLGen consortium. The summary statistics for other complex traits are available in publicly available resources, as listed in Table S5.

Additional data related to this paper may be requested from the authors.

