## Supplemental Material for "Mapping of the gene network that regulates glycan clock of ageing"

Azra Frkatović-Hodžić *et al.*

This file contains Supplementary Methods, Supplementary Figures and Appendix Tables.

Additional Supplementary Tables (S1-S6) are available in a separate file.

Supplementary Methods

**Studied cohorts**

Recruitment of participants, sample collection, genotyping and phenotyping in the cohorts used in the galactosylation GWAS was performed by staff members at the University of Zagreb, Croatia, University of Split Medical School, Croatia, UK, United Kingdom, King's College London, UK, German Institute of Human Nutrition, Germany, University of Tartu, Estonia, Leiden University Medical Centre (LUMC), Netherlands, Helmholtz Zentrum München – German Research Center for Environmental Health, Germany, and the nine study centers of the German Chronic Kidney Disease (GCKD) study.

IgG N-glycan quantification was performed by Genos Glycoscience Research Laboratory, Zagreb, Croatia and LUMC, Leiden, Netherlands.

**TwinsUK**

TwinsUK is a national registry of 12,000 volunteer twins in the UK. The cohort consists of 83% female subjects with a nearly equal number of monozygotic (51%) and dizygotic (49%) twin pairs. With an aim to study the genetics of healthy ageing and complex diseases, a sample of 7000 twins was assessed for a range of clinical, biochemical, behavioural and socio-economic characteristics. Moreover, several omics' datasets for the TwinsUK dataset are available including genome-wide SNP data. The study participants provided informed consent and ethical approval was obtained for

academic and commercial use of the study^1^.

**The European Prospective Investigation into Cancer and Nutrition Study**

The European Prospective Investigation into Cancer and Nutrition (EPIC) -Potsdam is a prospective cohort study that includes 27548 participants recruited from the Potsdam population in Germany from 1994 to 1998^2^. Participants’ age at recruitment ranges between 35 and 65, and the number of male and female subjects is 10904 and 16644, respectively. Initial data collection consisted of anthropometric measurements and blood sample collection used for omics' data derivation^3^. Ethical approval was obtained from the ethics committee in Germany and all participants provided informed consent^2^.

**CROATIA Vis, CROATIA-Split and CROATIA-Korcula**

CROATIA-Vis, CROATIA-Korcula and CROATIA-Split cohorts were collected as part of the “10001 Dalmatians” study, a study of Croatian island isolates with participants from six Adriatic islands (Korčula, Vis, Lastovo, Susak, Rab, Mljet) and the city of Split. The aim of the study is to investigate genetic and environmental determinants in health and disease by using the advantage of genetically isolated populations. In the recruitment process, a total of 1008 participants aged 19-93 were recruited for the CROATIA-Vis cohort in villages of Vis and Komiza during 2003 and 2004, 1012 subjects aged 18-85 were recruited in 2009-2010 in the city of Split for CROATIA-Split cohort and data on CROATIA-Korcula subjects (aged 18-98) was collected from the island of Korcula, specifically from the town of Korcula and three villages including Lumbarda, Zrnovo and Racisce. Participants were assessed for a number of anthropometric and physiological measurements, and they donated overnight fasting blood samples which were later used for DNA analysis, biochemical measurements and molecular marker assessment^4^. The study participants provided signed informed consent. Ethical approval was obtained for each cohort from ethics committees in Croatia and Scotland.

**The Orkney Complex Disease Study**

The Orkney Complex Disease Study (ORCADES) is a family-based cohort collected with the goal of identifying genetic risk factors in complex diseases in the population of isolated Orkney Island in northern Scotland. The recruitment started in 2005 and lasted for six years during which 2080 participants were recruited. The subjects were included if they had at least two of their grandparents who were Orcadian. The initial data collection included cardiovascular measurements and fasting blood sample collection, followed by subsequent visits for cognitive function assessment, eye measurements and DEXA scans. The study was approved by ethics committees in Scotland and all participants gave informed consent^5^.

**Leiden Longevity Study**

Leiden Longevity Study (LLS) is a family-based cohort from the Dutch population which was intended for studies of human longevity. Nonagenarian siblings (individuals having a sibling older than 89 years for men and 91 years for women) and their offspring and offspring's spouses (serve as controls) were included in the study if they were European. Initial recruitment started in 2002 and ended in 2006 during which blood samples were collected for assessment of plasma parameters and genetic material extraction. A total of 3359 subjects were included: 944 long-lived proband siblings, 1671 offspring and 744 controls (offspring’s spouses). This study was approved by the ethics committee of LUMC, Netherlands. Signed informed consent was given by all participants^6^. In the current study, a subset of 1190 participants including only offspring and their spouses is used.

**The Cooperative Health Research in The Augsburg Region F4**

The Cooperative Health Research in The Augsburg Region (KORA) F4 is a population-based study that was conducted between 2006 and 2008 as a follow-up of the KORA S4 study^7^. Participants were randomly selected from the population registry in the Augsburg region and two other neighbouring counties in Germany. The data collection included standard medical and physical examinations. A total of 3080 participants (51% females) aged 32-86 years were recruited^8^. Ethical approval for the study was obtained from the Ethics committee of Bavarian Chamber of Physicians, Germany. All participants provided signed informed consent before entering the study.

**The Viking Health Study - Shetland**

The Viking Health Study - Shetland (VIKING) is an epidemiologic study initiated to explore genetic risk factors for complex diseases. The cohort consists of individuals from an isolated population of Shetland in northern Scotland and the main criteria for recruitment was to have at least two grandparents from Shetland. Between 2013 and 2015, 2105 participants were recruited. A large number of distant relatives makes the VIKING cohort fit for the identification of rare genetic variants influencing the disease risk^9^. During initial data collection, data on health-related phenotypes and environmental parameters were collected and participants donated a fasting blood sample.

**The Estonian Genome Center of the University of Tartu Biobank**

The Estonian Genome Center of the University of Tartu (EGCUT) Biobank is a volunteer-based cohort of 52 thousand adult subjects aged ≥ 18 from the Estonian population. The recruitment was conducted throughout Estonia via general practitioners’ offices and medical personnel during the 2002-2012 period. Besides completing a questionnaire on topics such as lifestyle, diet and clinical diagnostics, participants also donated blood samples. The cohort was utilized in the exploration of more than 200 traits including anthropometric traits, common and rare diseases, blood biochemistry, as well as lifestyle and personality traits. The data on study participants is being continuously updated via follow-up health checks using electronic health registries and re-examinations^10^.

**German Chronic Kidney Disease Study**

German Chronic Kidney Disease (GCKD) study is an ongoing prospective study of kidney disease patients who are under nephrologist care in Germany^11^. The current sample size of 5217 makes it one of the largest chronic kidney disease cohort in the world. The subject enrolment was undertaken between 2010 and 2012 via nephrologist practice and outpatient care units of nine study centers throughout different regions in Germany. The mean age of study subjects is 60 years, with 40% of the participants being female. The data collection includes collecting information on sociodemographic factors, medical and family history, as well as obtaining blood samples in a standardized manner, which we immediately processed and stored frozen in a central biobank until measurement of the glycans.

**Genotyping, genotype QC and genotype imputation**

Genotyping was performed using commercially available SNP genotyping arrays, followed by genotype calling in Illumina and Genome Browser software**.** Quality control (QC) of genotype data was performed to exclude SNPs and samples with low quality including removal of 1) SNPs with low call rate, 2) SNPs violating the assumptions of Hardy-Weinberg Equilibrium (HWE) and 3) SNPs with low minor allele frequency (MAF) < 1%. Details on SNP exclusion criteria for each cohort and imputation to Haplotype Reference Consortium (HRC)^12^ panel are shown in Appendix Table 8. The genotypes were mapped to Genome Reference Consortium GRCh37 (hg19) build.

**IgG N-glycome analysis**

**IgG N-glycan Quantification** **by Ultra-Performance Liquid Chromatography**

Ultra-performance liquid chromatography (UPLC) is used for quantification of glycan structures attached to Fc (constant region) and Fab (variable region) portions of IgG without the possibility to distinguish them. Detailed protocol for UPLC quantification of IgG N-glycans is published elsewhere^13^.

Briefly, IgG was isolated from blood plasma samples using Protein G plates (BIA Separations, Ajdovščina, Slovenia). After filtration, plates were extensively washed to remove unwanted proteins and IgG was released from protein G monoliths using 0.1 M formic acid. Eluates were collected in a 96-well plate and neutralized with neutralization buffer (1 M ammonium bicarbonate) to pH 7.0 to maintain the stability of IgG. IgG samples were dried and denatured using SDS detergent and incubated at 65°C for 10 minutes. N-glycans from IgG were released using recombinant N-glycosidase F followed by fluorescent labelling with 2-aminobenzamide (2-AB) dye. Hydrophilic interaction liquid chromatography (HILIC) based solid-phase extraction (SPE) was used to remove excess protein, reagents and fluorescent label. Fluorescently labelled N-glycans were separated by hydrophilic interaction UPLC on Waters Aquity UPLC H-class instrument (Waters, Milford, MA) with Waters bridged ethylene hybrid (BEH) glycan chromatography column. A linear gradient of 75 to 62% ACN in a 20-min analytical run was used to separate different glycan structures. The retention times for individual glycans were converted to glucose units based on hydrolysed and 2-AB labelled glucose oligomers which were used as external standards for calibration of the system. Data processing was done in two ways depending on the cohort, 1) automatic integration as described in Agakova *et al*.^14^ or 2) using Empower 3 software with an automated processing method with traditional integration algorithm, followed by manual correction of each chromatogram to maintain the same integration intervals in all samples. The resulting chromatograms were separated into 24 peaks where the amount of glycans was expressed as % of the total integrated area in the corresponding peak (GP1-GP24). Total separation of each glycan structure is not possible using the described method, thus resulting in multiple glycan structures being detected under ten peaks. Glycan structures in each peak are listed in Appendix Table 9.

**Glycan Quantification by Liquid Chromatography coupled with Mass Spectrometry**

The full name of the method is reverse-phase nano-liquid-chromatography-sheath-flow-electrospray-mass spectrometry (LC-ESI-MS) but in this study, we refer to it as LC-MS. The detailed protocol for analysis of IgG N-glycans using LC-MS is described in Selman *et al*^15^. Briefly, IgG was isolated by affinity chromatography binding to protein G 96-well plates (BIA Separation, Ajdovščina, Slovenia) and treated with trypsin overnight at 37ºC which allowed cleavage of IgG at specific amino acid sites. The cleavage by trypsin resulted in different glycopeptides due to the difference of amino acid sequence in different IgG subclasses, thereby enabling subclass-specific glycan measurements. IgG subclass separation was performed using the Ultimate 3000 HPLC system (Dionex Corporation, Sunnyvale, CA). The SPE trap column was conditioned with mobile phase A and samples were loaded and separated on Ascentis Express C18 nano-LC column (Supelco, Bellefonte, USA) conditioned with mobile phase A and 95% ACN. For detection of separated subclass-specific glycopeptides, the HPLC system was coupled to a Dionex Ultimate UV detector and interfaced to a quadrupole-TOF-MS mass spectrometer (Bruker Daltonics, Bremen, Germany) with a standard ESI source (Bruker Daltonics, Bremen, Germany) and a sheath-flow ESI sprayer (Agilent Technologies, Santa Clara, USA). The mass spectra were recorded in a range between 300 and 2000 m/z with two averages at the frequency of 1Hz. The analysis time for one sample was 16 minutes. The calibration of LCMS datasets was done internally using a list of known glycopeptides and datasets were exported to the open mzXML format by Bruker DataAnalysis 4.0 software, followed by alignment to a master dataset of a typical sample. In-house software “Xtractor2D” was used to extract pre-defined features such as peak maximum or peak area in specific retention time and mass windows. Relative intensities of subclass-specific glycopeptides were obtained by integrating and summing three isotopic peaks. The obtained intensities were then normalized to the total IgG subclass-specific glycopeptide intensities. IgG2 and IgG3 subclasses have the same tryptic glycopeptide moieties, thus not enabling the separation of the subclass-specific glycopeptides. Here, obtained measurements are simply referred to as IgG2/3. LC-MS quantification results in 50 values which refer to 20 glycans measured on IgG1, 20 glycans on IgG2/3 and 10 glycans on IgG4. All glycans measured on IgG4 are fucosylated structures since the nonfucosylated glycans are hard to distinguish from the glycans found on IgG1^16^. The list of glycans measured by LC-MS and their description is listed in Appendix Table 10.

**IgG Glycan Data harmonization**

Previously, there were no GWA meta-analyses of IgG N-glycan patterns using GWAS of both UPLC- and LC-MS-derived IgG N-glycan traits, therefore making it necessary to first assess the correlation of the data and methods which should be applied in pre-processing step to make UPLC and LC-MS glycan traits comparable. For this purpose, we used the CROATIA-Vis cohort (n=661) as both UPLC and LC-MS IgG N-glycan measurements are available in the same samples.

We aimed to combine IgG subclass information obtained from LC-MS in an appropriate manner to obtain information corresponding to total IgG glycome values measured by UPLC. Pre-processing of IgG glycome data consists of normalization of the data and batch correction to remove the effects of experimental variation. Normalization procedure is necessary to remove unwanted variation between the samples and allows quantitative comparison of the samples^17^.  We tested the following three normalization types: total area normalization, largest peak normalization and median quotient normalization, all of which can be applied using “glycanr”^18^ package in R software^19^. We tested different normalizations both across the total glycome and per IgG subclass in LC-MS data.

Due to varying laboratory conditions during IgG N-glycan measurement, it was necessary to perform batch correction to remove non-biological, experimental variation. The batch correction was performed using ComBat function in R package “sva”^20^. We first log-transform the data as ComBat function implements empirical Bayes method for batch correction^21^ which assumes a normal distribution of the data, followed by batch correction with ComBat() where the batch is denoted by the plate on which the samples were analysed, and lastly, exponential transformation of the values to the original scale.

We calculated derived traits from the initial traits for purpose of data harmonization and to enable a more straightforward interpretation of the GWAS results so that the discovered genomic loci can be directly linked to the addition of one or two galactose residues to the IgG N-glycan chain: agalactosylation (G0), monogalactosylation (G1) and digalactosylation (G2). Formulas for calculation are listed in Appendix Table 11.

Since LC-MS data quantifies glycans attached to different IgG subclasses, in order to calculate different galactosylaiton traits we also incorporated the approximation of the IgG glycan subclass response factor (RF) to represent the IgG subclass concentration relative to other subclasses. RF is defined as the ratio between the concentration of the analyte and instrument response to the analyte. Using unpublished in-house experimental data,the subclass-specific response factors were approximated as follows: IgG1 with RF=1, IgG2/IgG3 with RF=2 and IgG4 with RF=1. IgG subclasses are present in different quantities in human serum; hence we also incorporated relative concentrations of each IgG subclass in the calculation of derived traits. The following relative measurements were indicated in the literature: 66% for IgG1, 30% for IgG2/IgG3 and 4% for IgG4^22^. The subclass-specific glycan measurements were weighted by the corresponding concentration and response factor before trait calculation.

**Pre-processing of glycan data**

Glycan data was pre-processed centrally in Genos for all cohorts except the LLS cohort for which the glycan data was pre-processed by a colleague from Leiden University Medical Centre. It is important to note that glycan data for the CROATIA-Korcula cohort was obtained in three instances (2010, 2013 and 2017) and each dataset was pre-processed separately and treated as an individual cohort in downstream analysis. Additionally, the TwinsUK cohort was analysed in four separate batches. Due to differences in sample collection, batches 1 and 2 were treated as one dataset and batches 3 and 4 were treated as the second dataset.

Data points in the 99.9^th^ percentile were removed and considered as technical outliers. Next, based on the results of the previous data harmonization assessment, median quotient normalization was applied on both UPLC and LC-MS glycan data across 24 and 50 glycan measurements, respectively and the batch correction was applied. Prior to the genetic association test, galactosylation traits in all cohorts were transformed using rank-based inverse normal transformation (mean=0, standard deviation=1).

**Supplementary Figures**

**G0**

**G2**

**G1**

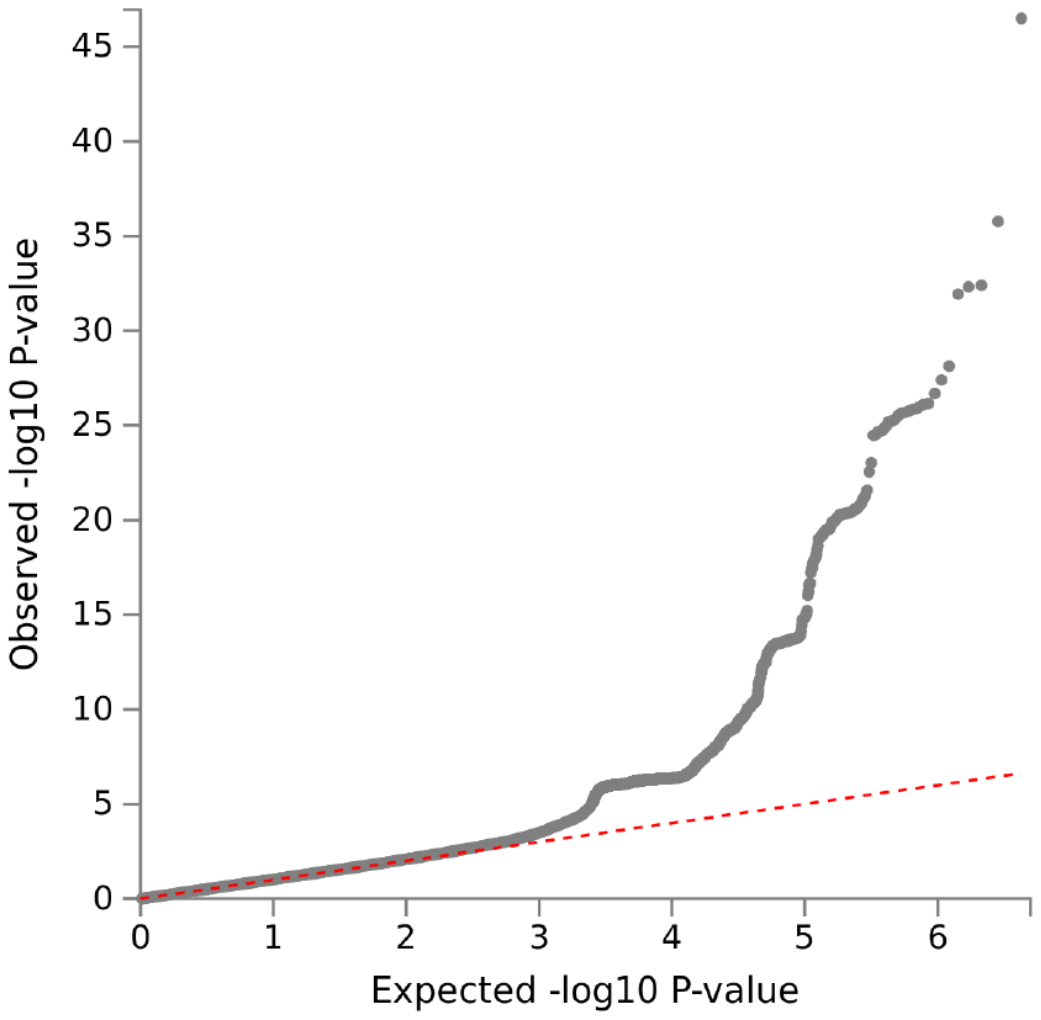

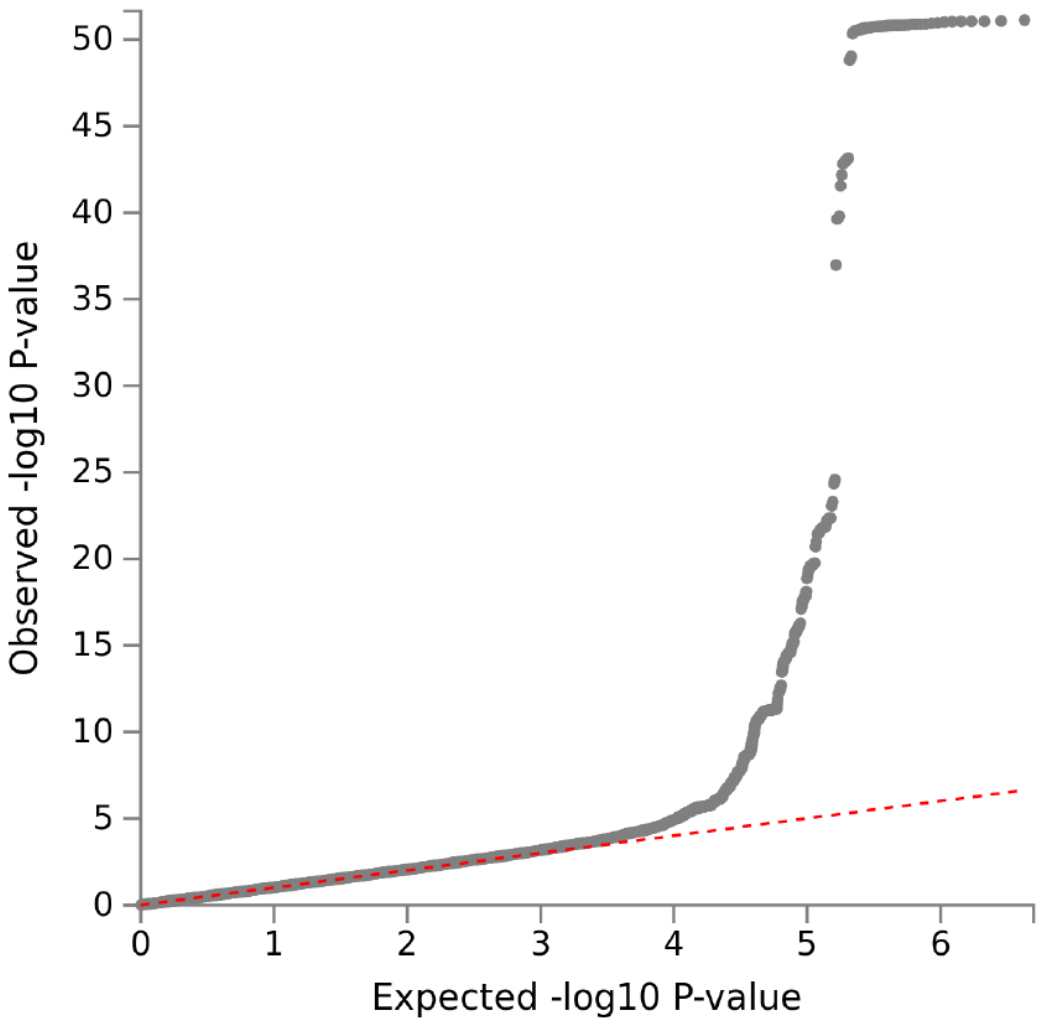

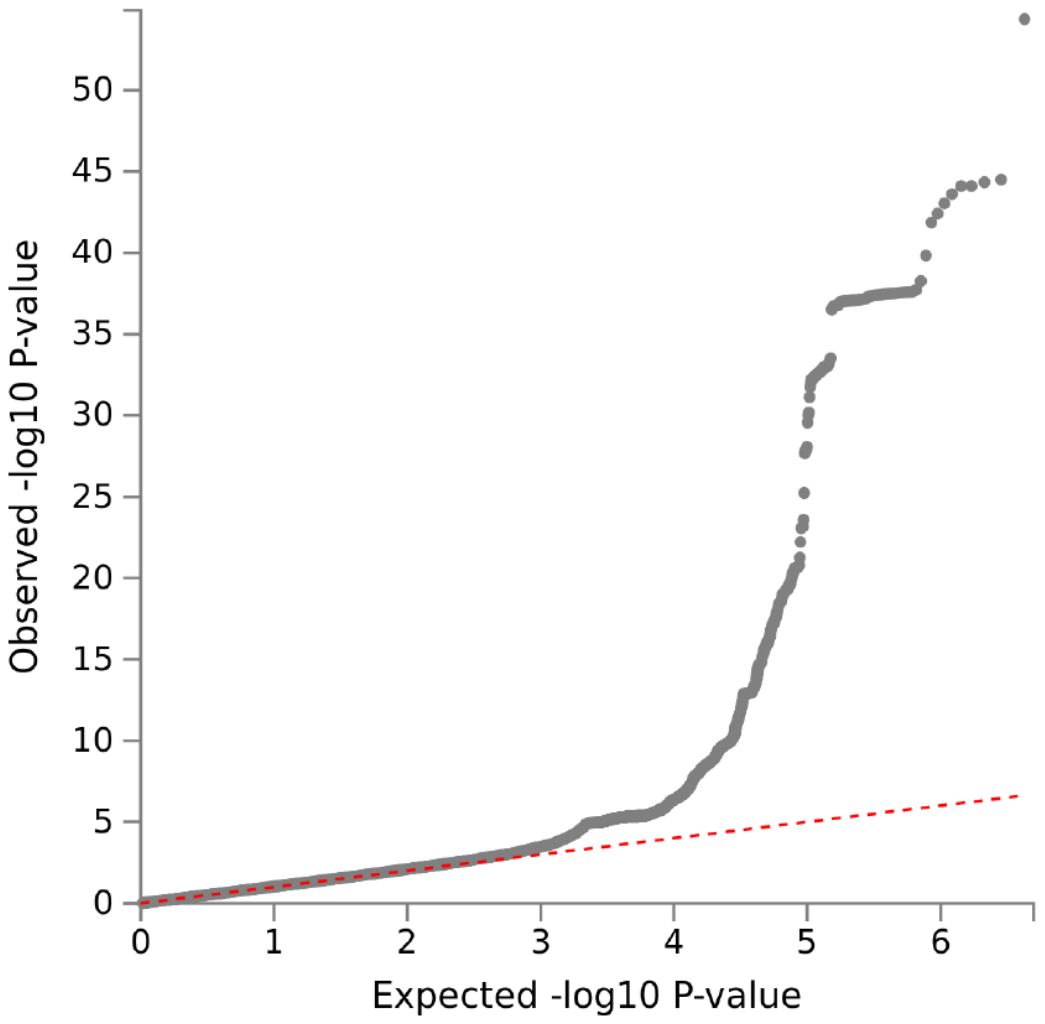

**Fig. S1**: QQ plots of association analysis for G0, G1 and G2 traits. Genomic inflation (lambda) for meta-analysis was 1.02, 1.01 and 1.03 for G0, G1 and G2, respectively.

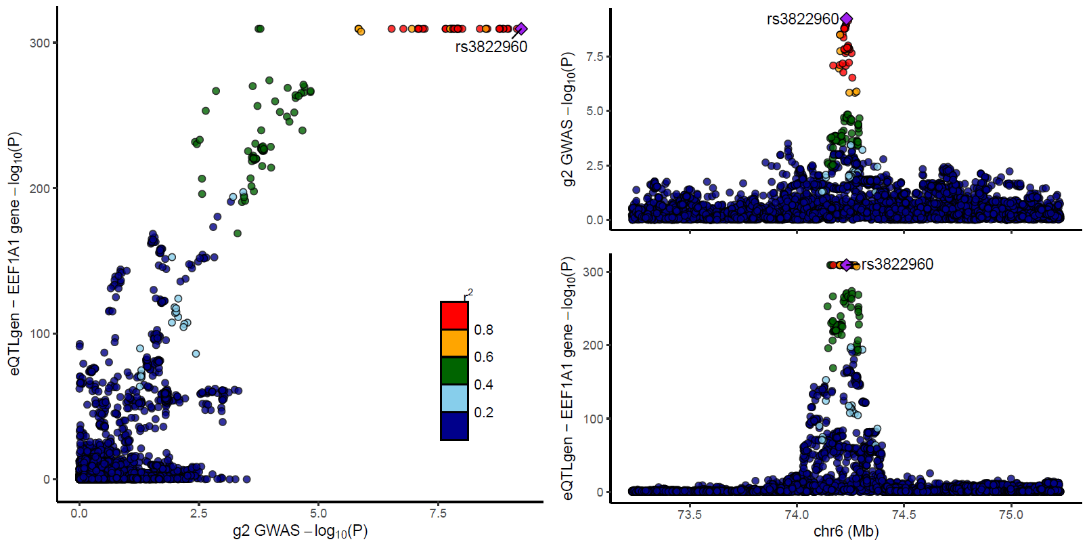

**a**

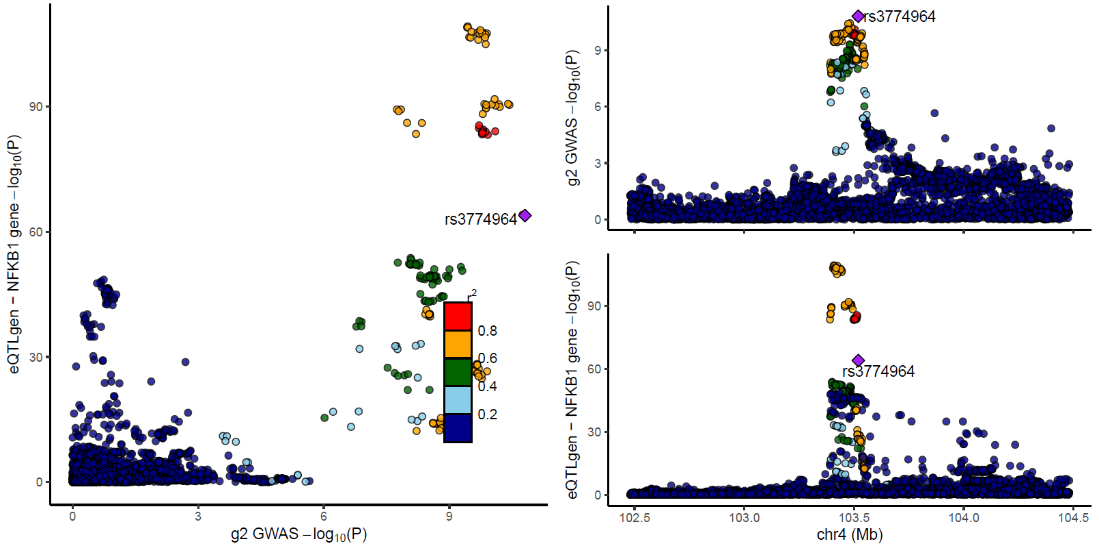

**b**

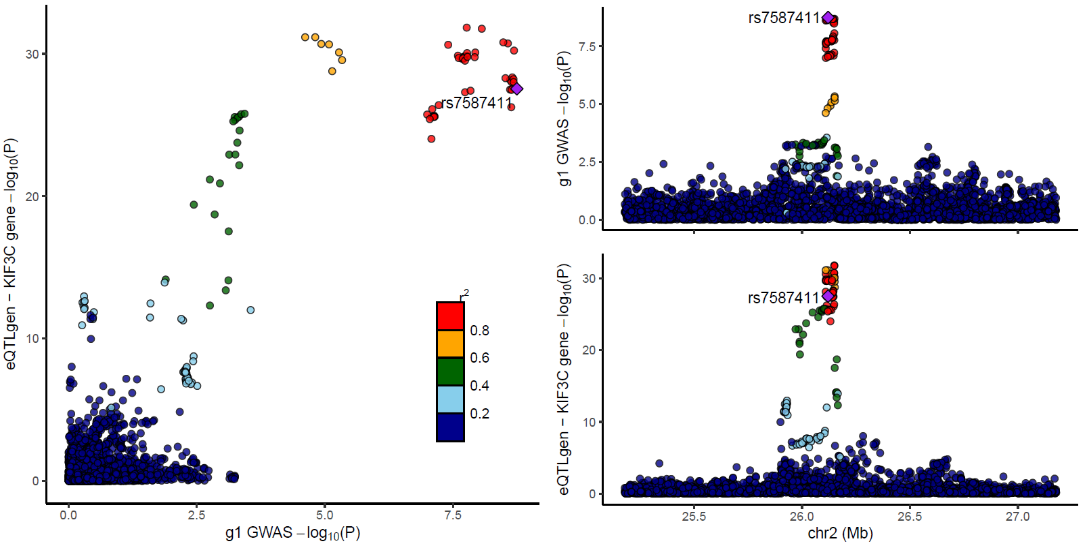

**c**

**Fig. S2.**: IgG galactosylation and eQTL association plots at loci with colocalization (PP4 > 75%). *Continued on next page*

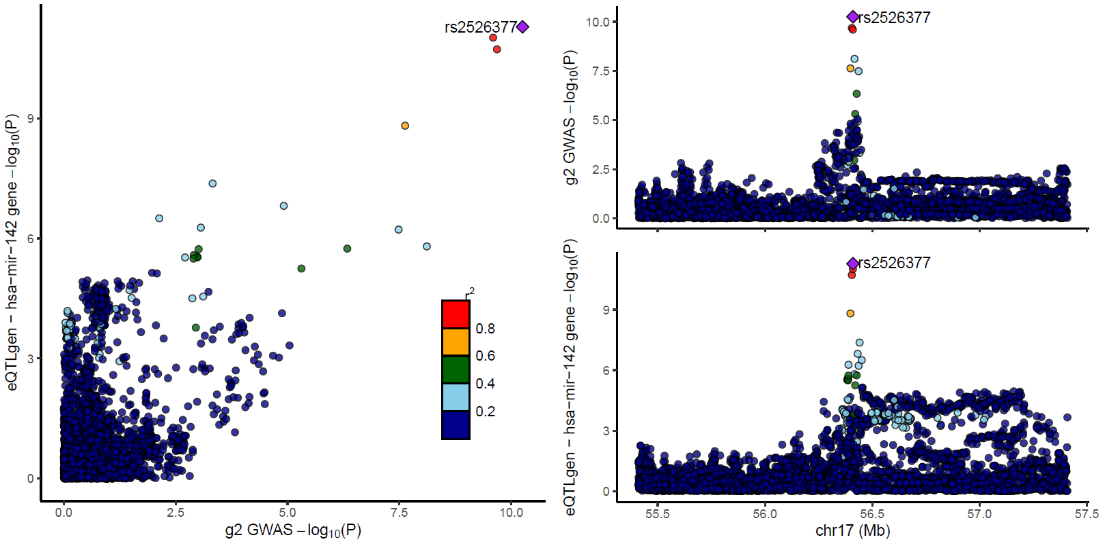

**d**

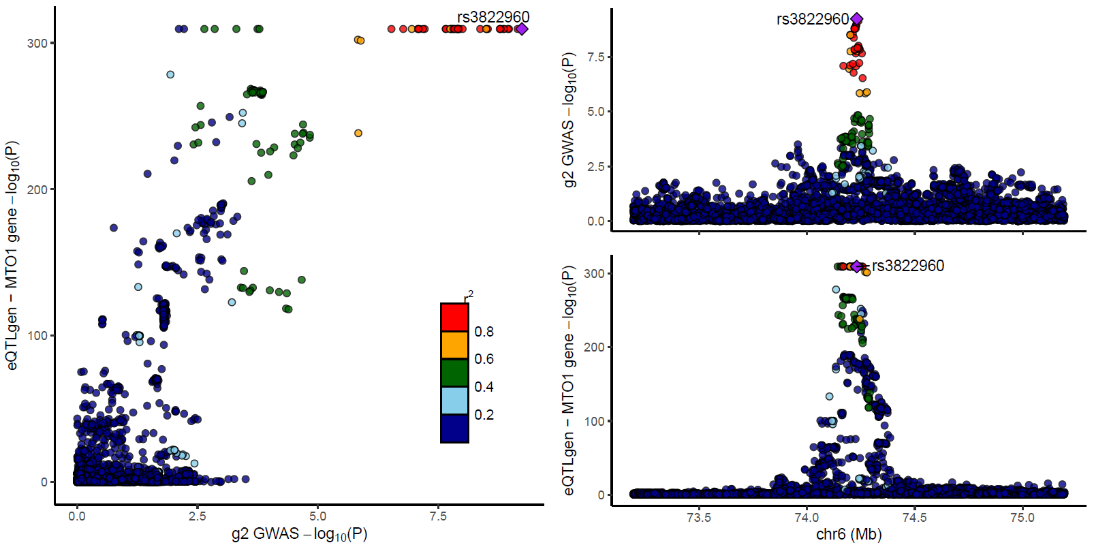

**e**

**Fig. S2**: IgG galactosylation and eQTL association plots at loci with colocalization (PP4 > 75%). Association plot for IgG galactosylation GWAS and eQTL are shown in the top right and bottom right corners, respectively; x-axis shows the chromosomal position, y-axis shows -log10(p-value) of the association. Correlation of -log10(p-values) for IgG galactosylation GWAS and eQTL are shown on left side. a) Colocalization in *EEF1A1* locus (prioritized genes *EEF1A1* and *MTO1*), b) Colocalization in *NFKB1* locus (prioritized genes *NFKB1* and *MANBA*), c) Colocalization in *KIF3C* locus (prioritized gene *KIF3C*), d) Colocalization in *hsa-mir-142* locus (prioritized genes *BZRAP1*, *SUPT4H1* and *RAD5C1*), e) Colocalization in *MTO1* locus (prioritized genes *EEF1A1* and *MTO1*).

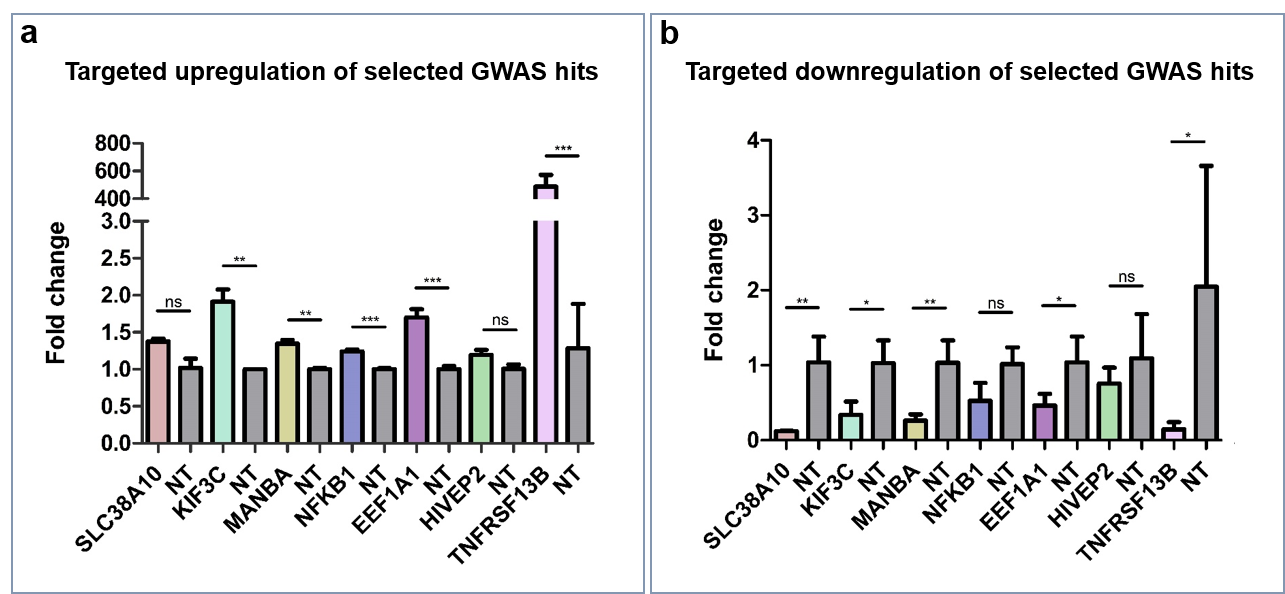

**Fig. S3.** Upregulation and downregulation of selected GWAS hits. Selected GWAS hits *associated* with IgG galactosylation (*SLC38A10, KIF3C, MANBA, NFKB1, EEF1A1, HIVEP2* and *TNFRSF13B*) were targeted in dCas9-VPR and dCas9-KRAB monoclonal cell lines. Samples containing non-targeting gRNAs served as controls. Changes of transcript levels are given as fold change values. **(a)** Targeting of selected GWAS hits with specific gRNA molecules in dCas9-VPR cell line resulted in significant increase of transcript levels of all loci except for *SLC38A10* and *HIVEP2*. **(b)** Targeting of the same genes in dCas9-KRAB call line resulted in significant downregulation of all genes except for *NFKB1* and *HIVEP2.* Statistical significance: *<0.05; **<0.01; ***<0.001; ns, not significant.

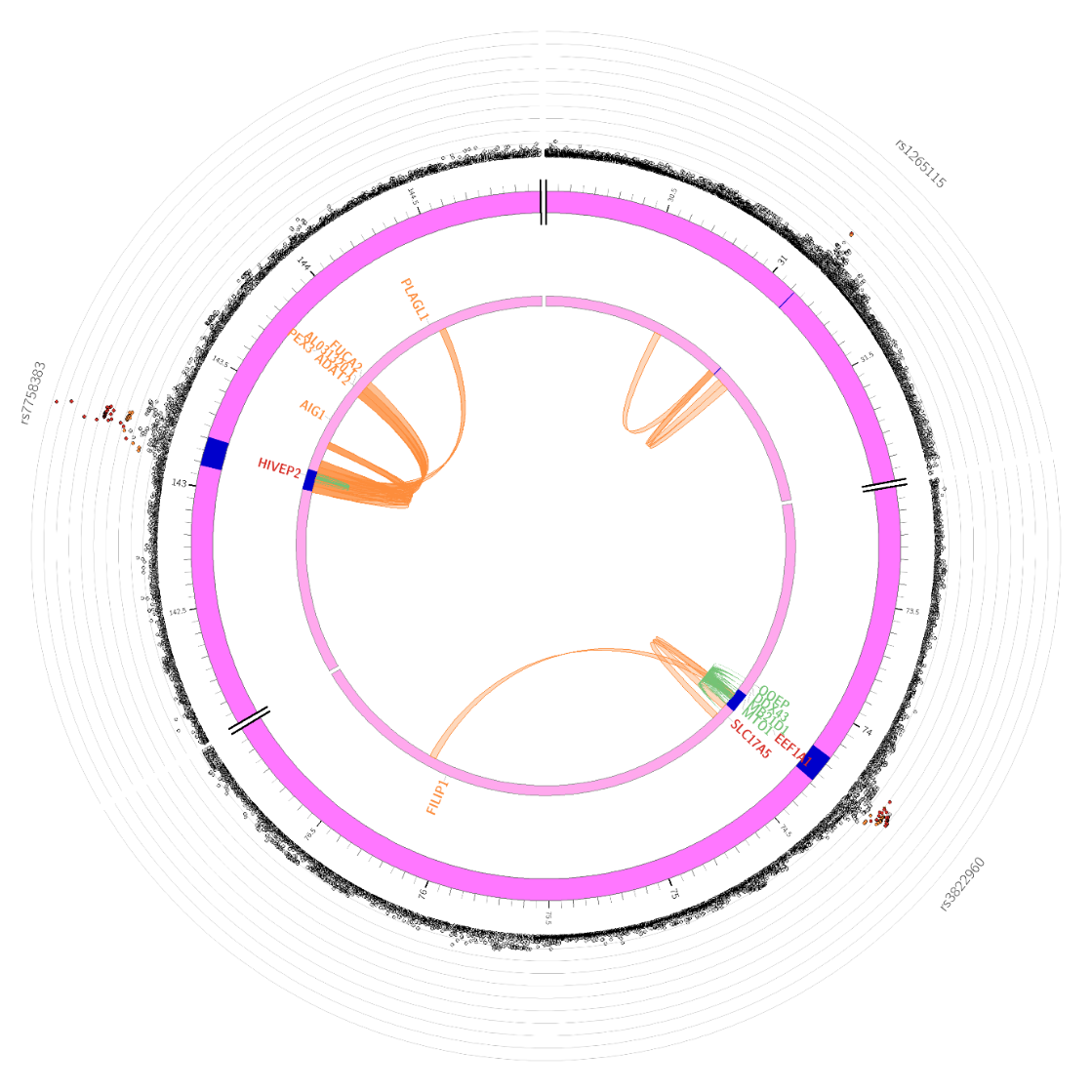

**Fig. S4**. Chromatin interaction map based on Hi-C data for GM12878 cell line (GSE87112) for chr6:143088071-143206826 locus indicating *FUCA2* and 4 other genes mapped via chromatin interaction with genetic variants in *HIVEP2* locus. Blue circle denotes the defined genomic locus; red circle denotes genes mapped via chromatin interaction mapping by FUMA.

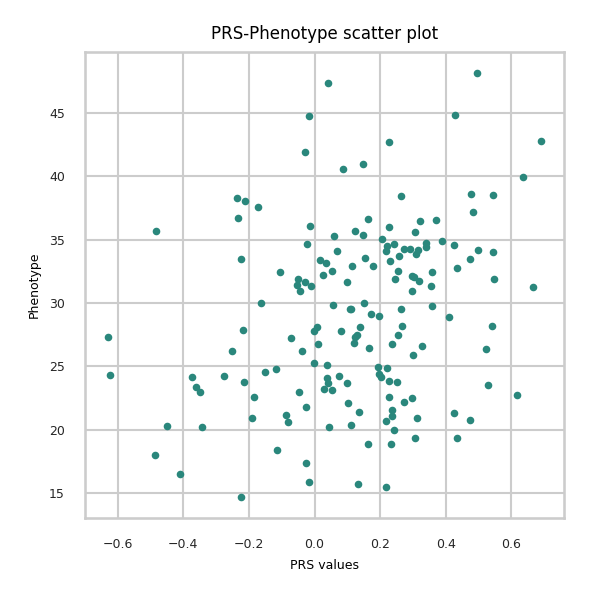

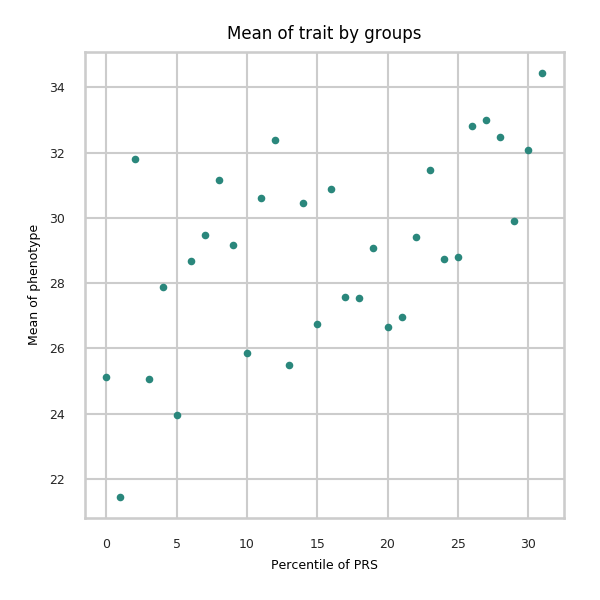

**Fig. S5.** PRS-Phenotype scatter plot (G0) and mean of trait by groups. The graph on the left-hand side shows the dependence of phenotype values (G0) on PRS values. The graph on the right-hand side is data stratified into 30 groups according to the PRS percentile. The Y-axis shows the average value of the trait for the group, the X-axis shows the group number.

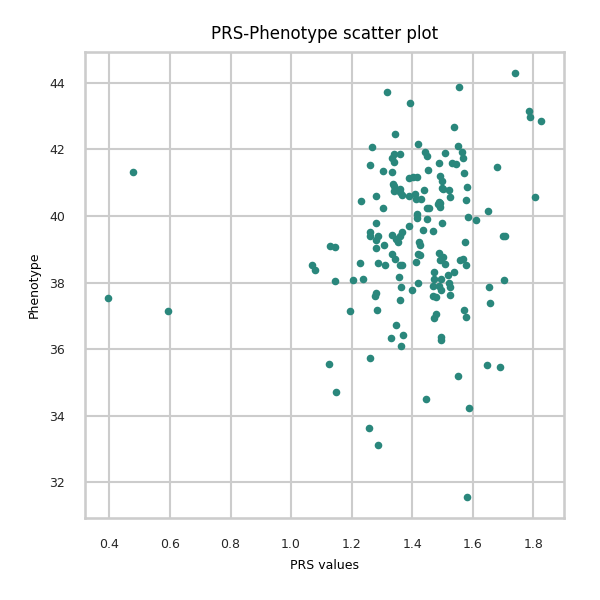

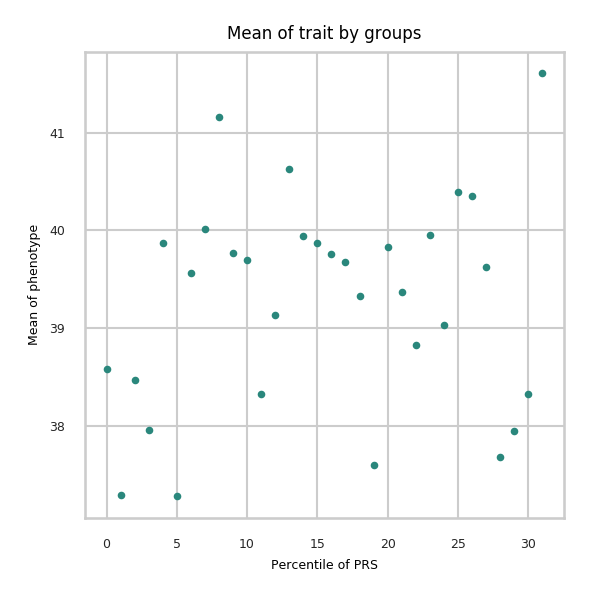

**Fig. S6**. PRS-Phenotype scatter plot (G1) and mean of trait by groups. The graph on the left-hand side shows the dependence of phenotype values (G1) on PRS values. The graph on the right-hand side shows data stratified into 30 groups according to the PRS percentile. The Y-axis shows the average value of the trait for the group, the X-axis shows the group number.

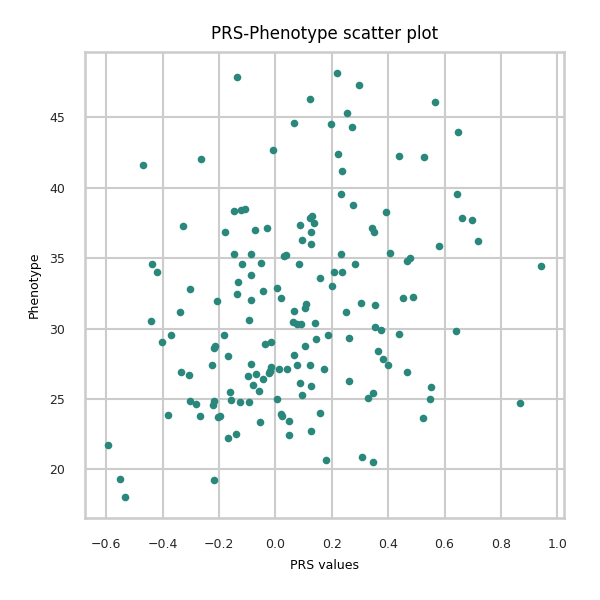

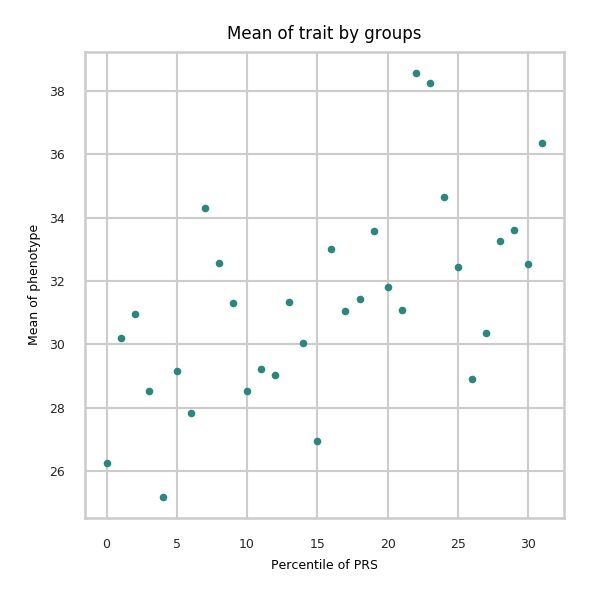

**Fig. S7**. PRS-Phenotype scatter plot (G2) and mean of trait by groups. The graph on the left-hand side shows the dependence of phenotype values (G2) on PRS values. The graph on the right-hand sideshows data stratified into 30 groups according to the PRS percentile. The Y-axis shows the average value of the trait for the group, the X-axis shows the group number.

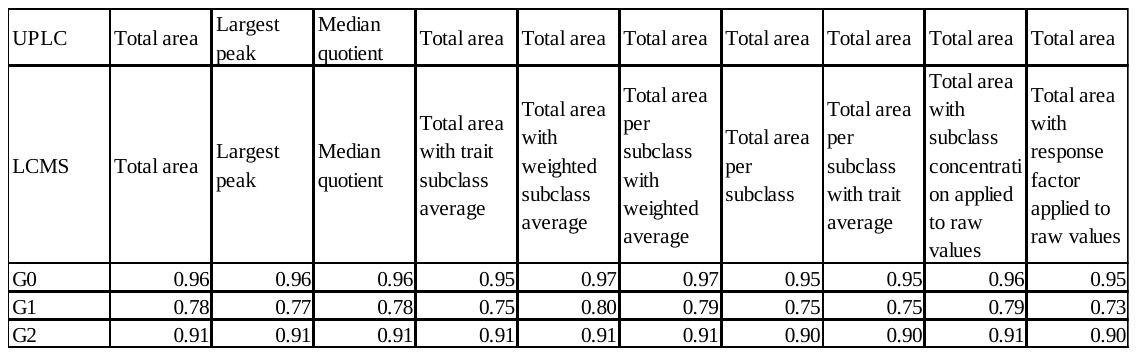

Appendix Table 1. Pearson's correlation coefficients for UPLC and LC-MS derived glycan traits after applying different normalization types and weighting of the subclass-specific values in LC-MS measurements. Combination of normalization types as applied in UPLC- and LC- MS-derived data is shown on the top.

| Discovery GWAS | | | | | | | | | | | | Replication GWAS | | | Previous GWAS | | | | | | |
| --- | --- | --- | --- | --- | --- | --- | --- | --- | --- | --- | --- | --- | --- | --- | --- | --- | --- | --- | --- | --- | --- |
| Locus | top SNP | top SNP chr | top SNP pos | Trait | EA | OA | EAF | Beta | StdErr | P-value | Beta repl | | SE repl | P repl | | Study | SNP | chr:pos | Trait | Pval | LD r2 with top SNP |
| 1:24699711-25493756 | rs188468174 | 1 | 25291697 | g0 | T | C | 0.015 | 0.693 | 0.048 | 3.20E-47 | 0.865 | | 0.076 | 2.96E-30 | | Klaric et al. | rs10903118 | 1:25294878 | IGP74 | 5.14E-13 | 0.01 |
| 2:26109539-26149988 | rs111919630 | 2 | 26139430 | g0 | T | C | 0.334 | -0.062 | 0.010 | 3.80E-10 | -0.049 | | 0.015 | 1.03E-03 | | Shadrina et al. | rs11895615 | 2:26113120 | Bisecting_GlcNac | 5.69E-10 | 0.87 |
| 4:103390496-103567348 | rs3774964 | 4 | 103519487 | g2 | A | G | 0.37 | -0.065 | 0.010 | 1.56E-11 | -0.038 | | 0.015 | 9.20E-03 | |  |  |  |  |  |  |
| 6:31107733-31164511 | rs1265109 | 6 | 31119589 | g0 | T | G | 0.295 | -0.063 | 0.011 | 1.57E-09 | 0.013 | | 0.019 | 4.97E-01 | | Klaric et al. | rs3099844 | 6:31448976 | IGP15 | 1.12E-13 | 0.02 |
| 6:74168723-74285118 | rs3822960 | 6 | 74230859 | g2 | T | C | 0.306 | 0.061 | 0.010 | 5.84E-10 | 0.028 | | 0.015 | 6.24E-02 | |  |  |  |  |  |  |
| 6:143088071-143206826 | rs7758383 | 6 | 143169723 | g2 | A | G | 0.486 | 0.092 | 0.009 | 6.09E-23 | 0.145 | | 0.014 | 9.44E-25 | | Klaric et al. | rs7758383 | 6:143169723 | IGP13 | 9.61E-14 |  |
| 7:150856165-150906453 | rs113745074 | 7 | 150942349 | g0 | T | C | 0.094 | 0.120 | 0.015 | 6.19E-16 | 0.134 | | 0.022 | 8.47E-10 | | Klaric et al. | rs7812088 | 7:150919829 | IGP2 | 2.06E-22 | 0.95 |
| 8:103542538-103550211 | rs13250010 | 8 | 103545983 | g0 | T | G | 0.356 | 0.063 | 0.010 | 7.71E-11 | 0.062 | | 0.015 | 3.47E-05 | | Klaric et al. | rs10096810 | 8:103545436 | IGP77 | 9.52E-11 | 0.9 |
| 9:32933492-33385427 | rs13297246 | 9 | 33128617 | g2 | A | G | 0.184 | 0.195 | 0.013 | 4.21E-55 | 0.202 | | 0.019 | 2.36E-26 | | Klaric et al. | rs10813951 | 9:33128021 | IGP17 | 8.84E-34 | 0.07 |
| 11:65555524-65555524 | rs10896045 | 11 | 65555524 | g1 | A | G | 0.301 | 0.079 | 0.013 | 2.61E-09 | 0.079 | | 0.017 | 2.42E-06 | | Shadrina et al. | rs479844 | 11:65551957 | N-glycosylation | 1.97E-13 | 0.54 |
| 17:16813994-16875636 | rs34562254 | 17 | 16842991 | g1 | A | G | 0.106 | 0.166 | 0.019 | 1.48E-18 | 0.092 | | 0.024 | 9.95E-05 | | Shadrina et al. | rs4561508 | 17:16848750 | N-glycosylation | 1.38E-10 | 0.88 |
| 17:43856639-44863413 | rs199516 | 17 | 44856485 | g0 | T | C | 0.221 | 0.068 | 0.012 | 1.20E-08 | 0.058 | | 0.018 | 1.25E-03 | | Klaric et al. | rs199456 | 17:44797919 | IGP14 | 6.76E-14 | 0.94 |
| 17:45766846-45870129 | rs1808192 | 17 | 45794706 | g2 | A | G | 0.351 | 0.061 | 0.010 | 5.94E-10 | 0.051 | | 0.015 | 6.78E-04 | | Klaric et al. | rs11651000 | 17:45835278 | IGP59 | 2.66E-12 | 0.23 |
| 17:56404349-56418136 | rs2526377 | 17 | 56410041 | g2 | A | G | 0.463 | 0.062 | 0.009 | 5.59E-11 | 0.057 | | 0.015 | 8.05E-05 | |  |  |  |  |  |  |
| 17:79158040-79268562 | rs2659005 | 17 | 79218714 | g2 | T | C | 0.436 | 0.079 | 0.009 | 4.36E-17 | 0.097 | | 0.016 | 6.99E-10 | | Klaric et al. | rs2725391 | 17:79192430 | IGP24 | 9.91E-16 | 0.74 |
| 21:36564553-36665202 | rs4817708 | 21 | 36567726 | g0 | T | C | 0.229 | 0.073 | 0.011 | 2.39E-11 | 0.043 | | 0.017 | 1.15E-02 | | Klaric et al. | rs7281587 | 21:36565278 | IGP45 | 1.13E-13 | 0.9 |

Appendix Table 2. List of IgG galactosylation associated loci and comparison of GWAS summary statistics for significantly associated genomic regions in current GWAS with previous GWAS of IgG N-glycome. Locus- chromosome: start-end position in GRCh37 (hg19) build; top SNP- rsID identifier for the top SNP in the locus; pos- chromosomal position of the top SNP; Trait- galactosylation trait with the strongest association with the locus;EA- effect allele; OA- non-effect allele; EAF- effect allele frequency; Beta- effect estimate for the top SNP; StdErr- standard error of the effect estimate; P-value- p value of the association; Study- Latest IgG N-glycome GWA study where the association was identified; SNP- rsID identified of the top SNP in the latest IgG N-glycome GWAS where the association was identified; chr:pos- chromosomal coordinates for the SNP in GRCh37 (hg19) build; Trait- trait with the strongest association with the SNP in the genomic region in GWAS denoted in Study column; Pval- p-value of the association; LD r2 with top SNP- LD r2 value between top SNP in discovery GWAS and top SNP in previous GWAS

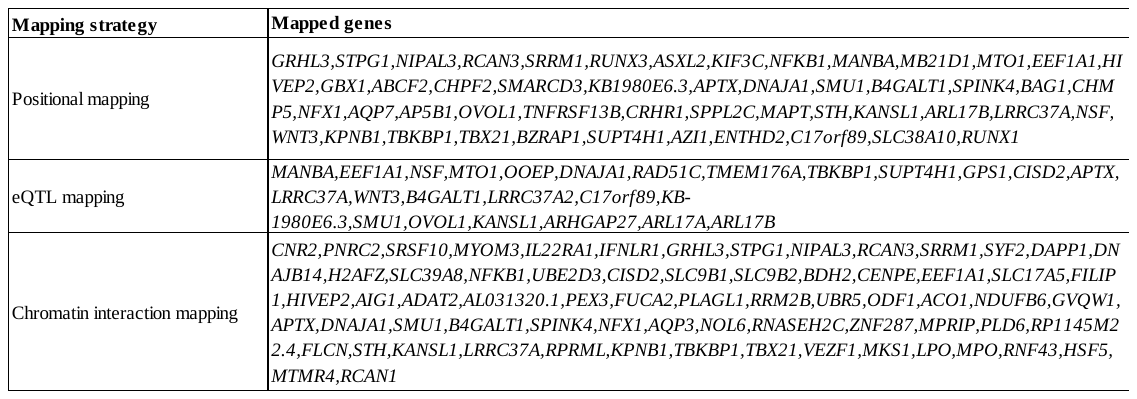

Appendix Table 3. List of genes mapped by positional, eQTL and chromatin interaction mapping in FUMA

| **Trait** | **Gene** | **Chr** | **Start** | **End** | **N SNPs** | **Z** | **P** | **P adjusted** |
| --- | --- | --- | --- | --- | --- | --- | --- | --- |
| g0 | RUNX3 | 1 | 25226002 | 25291612 | 113 | 4.207 | 1.29E-05 | 9.68E-03 |
| g2 | RUNX3 | 1 | 25226002 | 25291612 | 113 | 4.109 | 1.99E-05 | 1.33E-02 |
| g0 | NFKB1 | 4 | 103422486 | 103538459 | 276 | 6.163 | 3.57E-10 | 1.33E-06 |
| g0 | MANBA | 4 | 103552660 | 103682151 | 235 | 4.202 | 1.33E-05 | 9.77E-03 |
| g2 | NFKB1 | 4 | 103422486 | 103538459 | 276 | 6.506 | 3.86E-11 | 2.70E-07 |
| g2 | MANBA | 4 | 103552660 | 103682151 | 235 | 4.749 | 1.02E-06 | 1.24E-03 |
| g0 | EEF1A1 | 6 | 74225473 | 74233520 | 19 | 4.370 | 6.21E-06 | 5.61E-03 |
| g0 | HIVEP2 | 6 | 143072604 | 143266338 | 486 | 4.459 | 4.11E-06 | 4.05E-03 |
| g1 | HIVEP2 | 6 | 143072604 | 143266338 | 485 | 7.134 | 4.86E-13 | 9.07E-09 |
| g2 | MTO1 | 6 | 74171301 | 74218959 | 118 | 4.824 | 7.05E-07 | 9.39E-04 |
| g2 | EEF1A1 | 6 | 74225473 | 74233520 | 19 | 5.809 | 3.14E-09 | 8.37E-06 |
| g2 | HIVEP2 | 6 | 143072604 | 143266338 | 486 | 6.109 | 5.00E-10 | 1.65E-06 |
| g0 | ABCF2 | 7 | 150904923 | 150924316 | 70 | 5.770 | 3.97E-09 | 1.01E-05 |
| g0 | CHPF2 | 7 | 150929575 | 150935908 | 16 | 4.928 | 4.16E-07 | 6.30E-04 |
| g0 | SMARCD3 | 7 | 150935850 | 150974982 | 113 | 6.517 | 3.58E-11 | 2.70E-07 |
| g2 | ABCF2 | 7 | 150904923 | 150924316 | 70 | 4.621 | 1.91E-06 | 2.14E-03 |
| g2 | CHPF2 | 7 | 150929575 | 150935908 | 16 | 3.890 | 5.02E-05 | 2.84E-02 |
| g2 | SMARCD3 | 7 | 150935850 | 150974982 | 113 | 5.292 | 6.06E-08 | 1.13E-04 |
| g0 | KB-1980E6.3 | 8 | 103540968 | 103550896 | 60 | 6.235 | 2.25E-10 | 9.55E-07 |
| g2 | KB-1980E6.3 | 8 | 103540968 | 103550896 | 60 | 5.039 | 2.34E-07 | 3.86E-04 |
| g0 | DNAJA1 | 9 | 33025209 | 33039905 | 55 | 3.791 | 7.49E-05 | 3.99E-02 |
| g0 | SMU1 | 9 | 33041762 | 33076665 | 103 | 4.738 | 1.08E-06 | 1.28E-03 |
| g0 | B4GALT1 | 9 | 33104080 | 33167354 | 238 | 6.800 | 5.22E-12 | 4.87E-08 |
| g0 | SPINK4 | 9 | 33218363 | 33248565 | 103 | 5.628 | 9.11E-09 | 2.22E-05 |
| g0 | BAG1 | 9 | 33247818 | 33264761 | 22 | 4.264 | 1.01E-05 | 7.94E-03 |
| g0 | CHMP5 | 9 | 33264940 | 33281977 | 21 | 5.408 | 3.20E-08 | 6.39E-05 |
| g0 | NFX1 | 9 | 33290509 | 33371155 | 115 | 6.090 | 5.64E-10 | 1.76E-06 |
| g1 | B4GALT1 | 9 | 33104080 | 33167354 | 239 | 7.576 | 1.79E-14 | 5.00E-10 |
| g2 | DNAJA1 | 9 | 33025209 | 33039905 | 55 | 4.353 | 6.72E-06 | 5.85E-03 |
| g2 | SMU1 | 9 | 33041762 | 33076665 | 103 | 5.461 | 2.37E-08 | 4.91E-05 |
| g2 | B4GALT1 | 9 | 33104080 | 33167354 | 239 | 6.109 | 5.00E-10 | 1.65E-06 |
| g2 | SPINK4 | 9 | 33218363 | 33248565 | 103 | 6.352 | 1.06E-10 | 4.95E-07 |
| g2 | BAG1 | 9 | 33247818 | 33264761 | 22 | 4.350 | 6.79E-06 | 5.85E-03 |
| g2 | CHMP5 | 9 | 33264940 | 33281977 | 21 | 5.902 | 1.79E-09 | 5.01E-06 |
| g2 | NFX1 | 9 | 33290509 | 33371155 | 115 | 7.088 | 6.80E-13 | 9.52E-09 |
| g1 | OVOL1 | 11 | 65554493 | 65564690 | 25 | 4.144 | 1.71E-05 | 1.20E-02 |
| g0 | ARHGAP27 | 17 | 43471275 | 43511787 | 103 | 3.784 | 7.73E-05 | 4.08E-02 |
| g0 | PLEKHM1 | 17 | 43513266 | 43568115 | 97 | 3.762 | 8.44E-05 | 4.42E-02 |
| g0 | CRHR1 | 17 | 43699267 | 43913194 | 957 | 4.787 | 8.47E-07 | 1.09E-03 |
| g0 | SPPL2C | 17 | 43922256 | 43924438 | 18 | 4.712 | 1.23E-06 | 1.43E-03 |
| g0 | MAPT | 17 | 43971748 | 44105700 | 682 | 4.878 | 5.37E-07 | 7.70E-04 |
| g0 | STH | 17 | 44076616 | 44077060 | 1 | 4.884 | 5.20E-07 | 7.66E-04 |
| g0 | KANSL1 | 17 | 44107282 | 44302733 | 743 | 4.979 | 3.20E-07 | 5.11E-04 |
| g0 | ARL17B | 17 | 44352150 | 44439130 | 14 | 4.583 | 2.30E-06 | 2.43E-03 |
| g0 | NSF | 17 | 44668035 | 44834830 | 67 | 4.967 | 3.40E-07 | 5.29E-04 |
| g0 | WNT3 | 17 | 44839872 | 44910520 | 112 | 5.383 | 3.67E-08 | 7.09E-05 |
| g0 | TBX21 | 17 | 45810610 | 45823485 | 38 | 4.367 | 6.31E-06 | 5.61E-03 |
| g0 | AZI1 | 17 | 79163393 | 79196799 | 130 | 5.152 | 1.29E-07 | 2.33E-04 |
| g0 | ENTHD2 | 17 | 79202077 | 79212891 | 30 | 6.226 | 2.39E-10 | 9.55E-07 |
| g0 | C17orf89 | 17 | 79213039 | 79215081 | 6 | 5.998 | 9.98E-10 | 2.94E-06 |
| g0 | SLC38A10 | 17 | 79218800 | 79269347 | 185 | 5.490 | 2.01E-08 | 4.33E-05 |
| g1 | TNFRSF13B | 17 | 16832849 | 16875432 | 180 | 6.406 | 7.49E-11 | 4.20E-07 |
| g2 | CRHR1 | 17 | 43699267 | 43913194 | 957 | 4.294 | 8.78E-06 | 7.23E-03 |
| g2 | SPPL2C | 17 | 43922256 | 43924438 | 18 | 4.207 | 1.30E-05 | 9.68E-03 |
| g2 | MAPT | 17 | 43971748 | 44105700 | 682 | 4.394 | 5.57E-06 | 5.20E-03 |
| g2 | STH | 17 | 44076616 | 44077060 | 1 | 4.424 | 4.85E-06 | 4.61E-03 |
| g2 | KANSL1 | 17 | 44107282 | 44302733 | 743 | 4.515 | 3.16E-06 | 3.28E-03 |
| g2 | ARL17B | 17 | 44352150 | 44439130 | 14 | 4.182 | 1.44E-05 | 1.05E-02 |
| g2 | NSF | 17 | 44668035 | 44834830 | 67 | 4.608 | 2.03E-06 | 2.23E-03 |
| g2 | WNT3 | 17 | 44839872 | 44910520 | 112 | 5.056 | 2.14E-07 | 3.62E-04 |
| g2 | ITGB3 | 17 | 45331212 | 45421658 | 287 | 3.940 | 4.08E-05 | 2.38E-02 |
| g2 | ITGB3 | 17 | 45331263 | 45421658 | 287 | 3.940 | 4.08E-05 | 2.38E-02 |
| g2 | TBX21 | 17 | 45810610 | 45823485 | 38 | 4.771 | 9.15E-07 | 1.14E-03 |
| g2 | BZRAP1 | 17 | 56378592 | 56406152 | 82 | 4.075 | 2.30E-05 | 1.47E-02 |
| g2 | SUPT4H1 | 17 | 56422539 | 56430454 | 22 | 4.088 | 2.18E-05 | 1.42E-02 |
| g2 | AZI1 | 17 | 79163393 | 79196799 | 130 | 6.462 | 5.18E-11 | 3.22E-07 |
| g2 | ENTHD2 | 17 | 79202077 | 79212891 | 30 | 7.584 | 1.68E-14 | 5.00E-10 |
| g2 | C17orf89 | 17 | 79213039 | 79215081 | 6 | 6.366 | 9.68E-11 | 4.93E-07 |
| g2 | SLC38A10 | 17 | 79218800 | 79269347 | 185 | 7.012 | 1.18E-12 | 1.32E-08 |
| g0 | RUNX1 | 21 | 36160098 | 37376965 | 3508 | 4.784 | 8.60E-07 | 1.09E-03 |
| g2 | RUNX1 | 21 | 36160098 | 37376965 | 3506 | 4.440 | 4.51E-06 | 4.35E-03 |

Appendix Table 4. Results of genome-wide gene-based association test by MAGMA (FDR<0.05). Trait- galactosylation trait which was tested; Gene- symbol of the gene; Chr- chromosome on which gene is found; Start- starting position for the gene sequence in GRCh37(hg19) build; End- end position for the gene sequence; N SNPs- number of SNPs in the gene; Z- Z-score of the association; P- unadjusted P value of the association; P adjusted- adjusted p value using Benjamini & Hochberg method

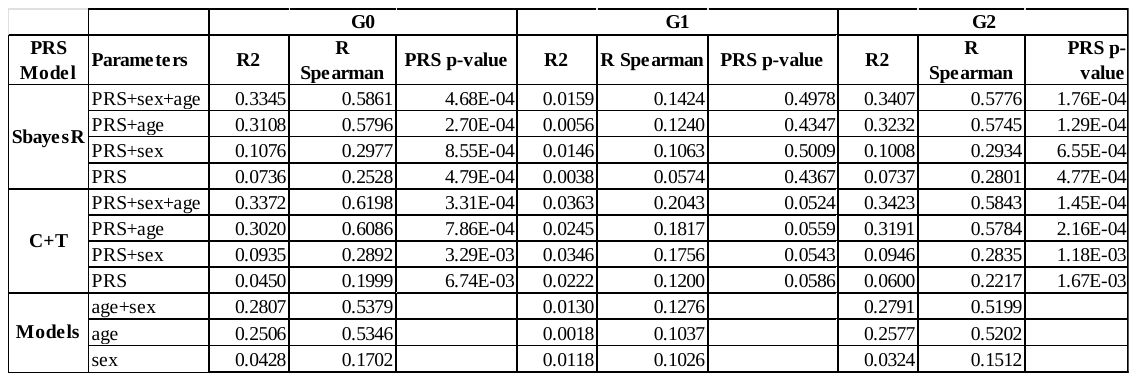

Appendix Table 5. Results of implementation of SBayesR and C+T models for G0, G1 and G2. PRS Model - method that was used to build model; Parameters - sets of parameters for a linear regression model with the trait as the outcome; R2 - part of trait variance that a model explains; R Spearman - the Spearman correlation coefficient between observed and predicted values of the trait; PRS p-value - the two-tailed p-value for the t-stats of PRS param.

| Experiment | Glycan trait | Estimate | SE | p-value | CI lower | CI upper |
| --- | --- | --- | --- | --- | --- | --- |
| VPR_EEF1A1 | GP_G0 | 2.484280 | 0.966877 | 0.010188 | 0.589236 | 4.379323 |
|  | GP_G1 | -2.246458 | 0.487330 | 0.000004 | -3.201608 | -1.291309 |
|  | GP_G2 | -0.257913 | 0.352768 | 0.464710 | -0.949325 | 0.433499 |
| VPR_HIVEP2 | GP_G0 | -1.408927 | 0.518379 | 0.006569 | -2.424931 | -0.392924 |
|  | GP_G1 | 0.117828 | 0.443605 | 0.790535 | -0.751621 | 0.987278 |
|  | GP_G2 | 1.079269 | 0.379913 | 0.004500 | 0.334654 | 1.823884 |
| VPR_MANBA | GP_G0 | 2.106936 | 1.137043 | 0.063883 | -0.121627 | 4.335500 |
|  | GP_G1 | -2.521960 | 0.755039 | 0.000837 | -4.001809 | -1.042112 |
|  | GP_G2 | -0.176833 | 0.265544 | 0.505459 | -0.972904 | 0.343625 |
| VPR_NFKB1 | GP_G0 | 0.823330 | 1.339071 | 0.538653 | -1.801201 | 3.447862 |
|  | GP_G1 | -0.886318 | 1.267960 | 0.484545 | -3.371473 | 1.598837 |
|  | GP_G2 | 0.022321 | 0.755465 | 0.976429 | -1.458363 | 1.503006 |
| VPR_TNFRSF13B | GP_G0 | 2.384531 | 0.622566 | 0.000128 | 1.164323 | 3.604739 |
|  | GP_G1 | -1.515340 | 0.902826 | 0.093261 | -3.284846 | 0.254166 |
|  | GP_G2 | -0.706994 | 0.360817 | 0.050063 | -1.414181 | 0.000194 |

Appendix Table 6. Meta-analysis results of testing differences in G0, G1 and G2 traits in treated (upregulation) and control cell line. Trait values were transformed to obtain normal distribution using Box-Cox method and analyzed using Student's t test. Estimate- effect size estimates; SE- standard error of estimate; p-value- p value of the meta-analysis; CI-confidence interval

| **Cohort** | **Platform for glycan analysis** | **Reference** | **N total available (glycans)** | **N female** | **N male** | **median age (min-max)** | **N in GWAS** |
| --- | --- | --- | --- | --- | --- | --- | --- |
| TwinsUK | UPLC | Menni *et al*. | 4624 | 4282 | 342 | 54 | 4477 |
| CROATIA-Korcula | UPLC and LC-MS | Pučić *et al*. | 2478 | 1148 | 1330 | 57 | 2436 |
| CROATIA-Split | LC-MS | Not published | 973 | 383 | 590 | 52 | 920 |
| CROATIA-Vis | LC-MS | Pučić *et al.* | 683 | 394 | 289 | 57 | 675 |
| VIKING | UPLC | Landini *et al.* | 1086 | 645 | 441 | 51 | 1071 |
| ORCADES | UPLC | Krištić *et al*. | 1786 | 1082 | 704 | 54 | 1720 |
| LLS | LC-MS | Ruhaak *et al.* | 1841 | 974 | 867 | 59 | 1190 |
| KORA F4 | LC-MS | Wahl *et al.* | 1823 | 935 | 888 | 62 | 1167 |
| EGCUT | UPLC | Trbojević-Akmačić *et al.* | 1108 | 516 | 592 | 69 | 483 |
| GCKD | UPLC | Not published | 4933 | 1965 | 2968 | 63 | 4933 |
| EPIC | UPLC | Not published | 3600 | 61.20% | 38.80% | 50 | 2406 |

Appendix Table 7. Overview of platforms used for quantification of IgG N-glycans and sample demographics. Cohort- name of the study; Platform- platform used for quantification of IgG N-glycans; Reference- published study in which the IgG N-glycome analysis for the cohort was described; N total (glycans)- number of subjects in the cohort with measured glycans; N female- number of female subjects; N male- number of male subjects; median age- median age of participants in the cohort; N in GWAS- number of pariticipants in GWAS

| **Cohort** | **Genotyping platform(s)** | **ID call rate** | **SNP call rate** | **HWE p** | **MAF** | **Imputation Tool** |
| --- | --- | --- | --- | --- | --- | --- |
| TwinsUK | Illumina HumanHap300; Illumina HumanHap610Q | >95% | > 97% (MAF ≥ 5%); > 99% (1% ≤ MAF < 5%) | >1e-6 | ≥1% | MACH (Michigan Imputation Server v1.0.2) |
| EPIC | Human660W-Quad_v1_A | >97% >99% | >95% | >1e-3 |  | Eagle2/minimac 3 |
|  | HumanCoreExome-12v1-0_B |  |  |  |  | Eagle2/minimac 3 |
|  | Illumina InfiniumOmniExpressExome-8v1-3_A DNA Analysis BeadChip |  |  |  | zCall threshold=7 | Eagle2/minimac 3 |
| LLS | Illumina660 W; Illumina OmniExpress | >95% | >95% | >1e-4 | ≥1% | IMPUTE2 |
| CROATIA-Korcula 1,2,3 | Illumina HumanHap s370CNV DUO/QUAD Phase 1(1); Illumina HumanOmniExpress Exome (2 & 3) | >97% | >98% | >1e-6 | ≥1% | SHAPEIT2/Sanger |
| CROATIA-Split | Illumina HumanHap 370CNV QUAD Phase I; Illumina HumanOmniExpress Exome | ≥97% | >98% | >1e-6 | ≥1% | SHAPEIT2/Sanger |
| CROATIA-Vis | Illumina HumanHap300v1 BeadChip | >97% | >98% | >1e-6 | ≥1% | SHAPEIT2/Sanger |
| VIKING | Illumina HumanOmniExpress Exome | >97% | >98% | >1e-6 | ≥1% omni markers; ≥0.01% exome markers | SHAPEIT2/Sanger |
| EGCUT | Illumina GSAv1.0, GSAv2.0, GSAv2.0_EST | >95% | >95% | >1e-4 | ≥1% | Beagle v.28Sep18.793 |
| KORA F4 | Affymetrix Axiom | >97% | >98% | >5e-6 | ≥1% | SHAPEIT/IMPUTE |
| ORCADES | HumanHap300v2 Phase 1 | >97% | >98% | >1e-6 | ≥1% | SHAPEIT2/Sanger |
| GCKD | Illumina Omni2.5Exome BeadChip | >97% | >96% | >1e-5 | >1% | Eagle/minimac3 |

Appendix Table 8. Overview of genotyping arrays and imputation. Cohort- name of the participating study; Genotyping platform- SNP array used for genotyping in the given cohort; ID call rate- ID call rate; SNP call rate- SNP call rate; HWE p- p value of HWE test used for filtering; MAF- minor allele frequency threshold used for filtering; Imputation tool- software used for imputation of genetic data;

|  | **Glycan name** | **Description** |
| --- | --- | --- |
| UPLC measured glycan traits | GP1 | FA1 glycan |
|  | GP2 | A2 glycan |
|  | GP3 | Structure not determined |
|  | GP4 | FA2 glycan |
|  | GP5 | M5 glycan |
|  | GP6 | FA2B glycan |
|  | GP7 | A2G1 glycan |
|  | GP8 | FA2[6]G1 glycan |
|  | GP9 | FA2[3]G1 glycan |
|  | GP10 | FA2[6]BG1 glycan |
|  | GP11 | FA2[3]BG1 glycan |
|  | GP12 | A2G2 glycan |
|  | GP13 | A2BG2 glycan |
|  | GP14 | FA2G2 glycan |
|  | GP15 | FA2BG2 glycan |
|  | GP16 | FA2G1S1 glycan |
|  | GP17 | A2G2S1 glycan |
|  | GP18 | FA2G2S1 glycan |
|  | GP19 | FA2BG2S1 glycan |
|  | GP20 | Structure not determined |
|  | GP21 | A2G2S2 glycan |
|  | GP22 | A2BG2S2 glycan |
|  | GP23 | FA2G2S2 glycan |
|  | GP24 | FA2BG2S2 glycan |

**Appendix Table 9.** Glycan names and description of the most abundant glycan structure in each peak as measured by ultra-performance liquid chromatography; Glycan name- name of the glycan; Description- strcture of the glycan (F- core fucosylated, A1- monoantennary, A2- diantennary, B- bisecting GlcNAc, G1- monogalactosylated, G2- digalactosylated, S1- monosialylated, S2- disialylated, M5- pentamannose, [3] or [6]- position of the attached galactose)

| **Glycan class** | **Glycan Name** | **Description** |
| --- | --- | --- |
| Total IgG1 glycans | LC_IGP1 | IgG1_G0 |
|  | LC_IGP2 | IgG1_G0F |
|  | LC_IGP3 | IgG1_G0FN |
|  | LC_IGP4 | IgG1_G0N |
|  | LC_IGP5 | IgG1_G1 |
|  | LC_IGP6 | IgG1_G1F |
|  | LC_IGP7 | IgG1_G1FN |
|  | LC_IGP8 | IgG1_G1FNS1 |
|  | LC_IGP9 | IgG1_G1FS1 |
|  | LC_IGP10 | IgG1_G1N |
|  | LC_IGP11 | IgG1_G1NS1 |
|  | LC_IGP12 | IgG1_G1S1 |
|  | LC_IGP13 | IgG1_G2 |
|  | LC_IGP14 | IgG1_G2F |
|  | LC_IGP15 | IgG1_G2FN |
|  | LC_IGP16 | IgG1_G2FNS1 |
|  | LC_IGP17 | IgG1_G2FS1 |
|  | LC_IGP18 | IgG1_G2N |
|  | LC_IGP19 | IgG1_G2NS1 |
|  | LC_IGP20 | IgG1_G2S1 |
| Total IgG2 glycans | LC_IGP87 | IgG2_G0 |
|  | LC_IGP88 | IgG2_G0F |
|  | LC_IGP89 | IgG2_G0FN |
|  | LC_IGP90 | IgG2_G0N |
|  | LC_IGP91 | IgG2_G1 |
|  | LC_IGP92 | IgG2_G1F |
|  | LC_IGP93 | IgG2_G1FN |
|  | LC_IGP94 | IgG2_G1FNS1 |
|  | LC_IGP95 | IgG2_G1FS1 |
|  | LC_IGP96 | IgG2_G1N |
|  | LC_IGP97 | IgG2_G1NS1 |
|  | LC_IGP98 | IgG2_G1S1 |
|  | LC_IGP99 | IgG2_G2 |
|  | LC_IGP100 | IgG2_G2F |
|  | LC_IGP101 | IgG2_G2FN |
|  | LC_IGP102 | IgG2_G2FNS1 |
|  | LC_IGP103 | IgG2_G2FS1 |
|  | LC_IGP104 | IgG2_G2N |
|  | LC_IGP105 | IgG2_G2NS1 |
|  | LC_IGP106 | IgG2_G2S |
| Total IgG4 glycans | LC_IGP173 | IgG4_G0F |
|  | LC_IGP174 | IgG4_G0FN |
|  | LC_IGP175 | IgG4_G1F |
|  | LC_IGP176 | IgG4_G1FN |
|  | LC_IGP177 | IgG4_G1FNS1 |
|  | LC_IGP178 | IgG4_G1FS1 |
|  | LC_IGP179 | IgG4_G2F |
|  | LC_IGP180 | IgG4_G2FN |
|  | LC_IGP181 | IgG4_G2FNS1 |
|  | LC_IGP182 | IgG4_G2FS1 |

Appendix Table 10. Glycan names and description of glycan structure in each peak as measured by liquid chromatography coupled with mass spectrometry; Glycan name- name of the glycan in LC-MS data; Description- structure of the glycan (F- core fucosylated, N- bisecting GlcNAc, G1- monogalactosylated, G2- digalactosylated, S1- monosialylated)

| **IgG N-glycan trait** | **Description** | **LCMS formula** | **UPLC formula** |
| --- | --- | --- | --- |
| G0 | Percentage of agalactosylated structures in total IgG N-glycome | (LC_IGP1+LC_IGP2+LC_IGP3+LC_IGP4+LC_IGP87+LC_IGP88+LC_IGP89+LC_IGP90+LC_IGP173+LC_IGP174)/SUM(ALL)*100 | SUM(GP1+GP2+GP4+GP5+GP6)/SUM(GP1:GP24)*100 |
| G1 | Percentage of monogalactosylated structures in total IgG N-glycome | (SUM(LC_IGP5:LC_IGP12)+SUM(LC_IGP91:LC_IGP98)+SUM(LC_IGP175:LC_IGP178))/SUM(ALL)*100 | (GP7+GP8+GP9+GP10+GP11+GP16)/SUM(GP1:GP24)*100 |
| G2 | Percentage of digalactosylated structures in total IgG N-glycome | (SUM(LC_IGP13:LC_IGP20)+SUM(LC_IGP99:LC_IGP106)+SUM(LC_IGP179:LC_IGP182))/SUM(ALL)*100 | (GP12+GP13+GP14+GP15+GP17+GP18+GP19+GP21+GP22+GP23+GP24)/SUM(GP1:GP24)*100 |

Appendix Table 11. Formulas used for calculation of G0, G1 and G2 traits from LC-MS and UPLC measured glycan data

| --pi | 0.95,0.02,0.02,0.01 |
| --- | --- |
| --gamma | 0.0,0.01,0.1,1 |
| --hsq | 0.5 |
| --chain-length | 3000 |
| --burn-in | 1000 |
| --unscale-genotype | yes |
| --exclude-mhc | yes |
| --robust | yes |
| --p-value | 0.9 |

Appendix Table 12. Parameters for SBayesR model for PRS calculation

| **sgRNA molecule** | **Target site sequence (5'→3')** | **Experiment** |
| --- | --- | --- |
| KIF3C_A-g1 | GTGTCCGAGGATGGTGGCTCG | CRISPRa (dSaCas9) |
| KIF3C_A-g2 | GTTCCCGGCATGCTCGGGCTCG |  |
| KIF3C_A-g3 | GTTGTAGTCCTGCCTTTGTTAA |  |
| NFKB1_A-g1 | GTATGCTCTCTCGACGTCAGT |  |
| NFKB1_A-g2 | GTCGAGAGAGCATACAGACAG |  |
| NFKB1_A-g3 | GGGCGGGAGGTAGGGTCGCTG |  |
| MANBA_A-g1 | GACCCGCGACGCCCGGCTTTCA |  |
| MANBA_A-g2 | GAGCGCTCAGCTGACCTAGGG |  |
| MANBA_A-g3 | GTCAGTACATACAACTGGAGGC |  |
| SLC38A10_A-g1 | GGTTTGGTGGGACGCGAGCCA |  |
| SLC38A10_A-g2 | GTCCAAACCGAGGCCTCGCGGG |  |
| SLC38A10_A-g3 | GCCAGCTCGTTGAACTGGGGCC |  |
| TNFRSF13B_A-g1 | GTGTGTACCCCATGGGACGCC |  |
| TNFRSF13B_A-g2 | GTTAGTGCTTGGGCTGCACTTC |  |
| TNFRSF13B_A-g3 | GCTTGCGATGAGGTATTGACTA |  |
| EEF1A1_A-g1 | GCCTCAGTGATGACGAGAGCGG |  |
| EEF1A1_A-g2 | GTGAGGCTCCGGTGCCCGTCA |  |
| EEF1A1_A-g3 | GTCGGCAATTGAACCGGTGCC |  |
| HIVEP2_A-g1 | GTCATCTGCATAGATCCGCGGC |  |
| HIVEP2_A-g2 | GGAGCTTGTTTCACTTTATAT |  |
| HIVEP2_A-g3 | GCTGCTTTGCGTAATGTCCTGG |  |
| KIF3C_G-g1 | GCCAGCTAAGCCTCCCGACTG | CRISPRi (dSpCas9) |
| KIF3C_G-g2 | GCGGCATGCTCGGGCTCGGTG |  |
| KIF3C_G-g3 | GCCTGTCATCCCGGAGCGGT |  |
| NFKB1_G-g1 | GGAAGAGGAGGTTTCGCCAC |  |
| NFKB1_G-g2 | GGCGGCCCGACCGACGGGAA |  |
| NFKB1_G-g3 | GCGCCGCCTTACTGGCCGGG |  |
| MANBA_G-g1 | GCTGTAACTGAGCTCCGCGG |  |
| MANBA_G-g2 | GTCGATCTCTCCACATCTCGG |  |
| MANBA_G-g3 | GCCTGATCCAGGTATGGCGTG |  |
| SLC38A10_G-g1 | GTCCCACCAAACCCTACCCCC |  |
| SLC38A10_G-g2 | GGTCACAGGTCCCAACGTCC |  |
| SLC38A10_G-g3 | GGTGTCCCTAGCTGGCTCTG |  |
| TNFRSF13B_G-g1 | GCTGGACCTTGCAACCCCCA |  |
| TNFRSF13B_G-g2 | GCCCAGGCCACTCATTACTC |  |
| TNFRSF13B_G-g3 | GCGAGGTGGCCGGAGCCGTG |  |
| EEF1A1_G-g1 | GGTGCCACCAGATTCGCACG |  |
| EEF1A1_G-g2 | GTTGCGAAAAAGAACGTTCA |  |
| EEF1A1_G-g3 | GAAGGGCCATAACCCGTAAAG |  |
| HIVEP2_G-g1 | GCTTTGCGTAATGTCCTGGG |  |
| HIVEP2_G-g2 | GGCCCGTTGTCAGGTAGCGG |  |
| HIVEP2_G-g3 | GCGACGTGCACCGGCGCGCA |  |
| NT-sgRNA | GTAGGCGCGCCGCTCTCTAC | Non-targeting control |

Appendix Table 13. gRNA sequences for targeted upregulation/downregulation

| **Primer name** | **Sequence (5'→3')** |
| --- | --- |
| KIF3C_FW1 | CATGGAGAGGCTCATGCGAT |
| KIF3C_REV1 | TGGTTGTCACTCATGGTCCG |
| NFKB1_FW1 | GAAGCACGAATGACAGAGGC |
| NFKB1_REV1 | GCTTGGCGGATTAGCTCTTTT |
| MANBA_FW1 | TGAGCTGCGTTTCCAGTCAG |
| MANBA_REV1 | ACATGGCATTCACCCTTCTGC |
| SLC38A10_FW1 | GGTCCCAACACAGGCTACTG |
| SLC38A10_REV1 | GACACCAAACAGCATCCACC |
| TNFRSF13B_FW1 | GCAAGGAGCAAGGCAAGTTC |
| TNFRSF13B_REV1 | TCCTTGGTACCTTCCCGAGT |
| EEF1A1_FW1 | TTGGACACGTAGATTCGGGC |
| EEF1A1_REV1 | CGTTCACGCTCAGCTTTCAG |
| HIVEP2_FW1 | GGGACACAGCTCTAGGACAAA |
| HIVEP2_REV1 | TGCGGTGGATATTGCTTCTCT |

Appendix Table 14. qPCR primer sequences for SYBR detection of gene expression.
